# Integrative analysis of blood cells DNA methylation, transcriptomics and genomics identifies novel epigenetic regulatory mechanisms of insulin resistance during puberty in children with obesity

**DOI:** 10.1101/2022.12.13.22283415

**Authors:** A Anguita-Ruiz, A Torres-Martos, FJ Ruiz-Ojeda, J Alcalá-Fdez, G Bueno, M Gil-Campos, J Roa-Rivas, LA Moreno, A Gil, R Leis, CM Aguilera

**Author notes:** **Corresponding Author:** Concepción M. Aguilera Instituto de Nutrición y Tecnología de los Alimentos, Instituto de Investigación Biomédica, Universidad de Granada. Parque tecnológico de la Salud, 18010 Armilla; Granada, Spain.

## Abstract

**Background:** Puberty is a time of considerable metabolic and hormonal changes associated with a physiological increase in peripheral tissue insulin resistance (IR). There is evidence that physiological IR does not resolve in youth who are obese, which may result in increased cardio-metabolic risk. Understanding the molecular and biological processes underlying the development of IR in puberty and the additional impact of obesity on these processes is crucial to prevent type 2 diabetes.

**Methods:** This is a longitudinal study based on the follow-up until puberty of a cohort of prepubertal Spanish boys and girls. The study population was composed of 139 children organized in a longitudinal approach of 90 subjects (47 females) and two cross-sectional approaches of 99 (52 females) and 130 (71 females) subjects for prepubertal and pubertal stages, respectively. Children were allocated into experimental groups according to their obesity and IR status before and after the onset of puberty. All participants presented blood DNA samples for GWAS and EWAS analyses. In 44 children of the pubertal stage, we counted on blood RNA samples for RNA-seq analysis.

**Results:** Our large-scale integrative molecular analysis identified novel blood multi-omics signatures (mapping the loci *ABCG1, ESR1* and *VASN*, among others) significantly associated with IR longitudinal trajectories in children with obesity during pubertal maturation. Functional enrichment analysis revealed that identified loci participate in systemic metabolic pathways and sexual maturation processes relevant to the pathogenesis of IR in the context of puberty. Additional analyses on cardiometabolic and inflammatory phenotypes showed that blood DNAm patterns of some of the identified loci are further associated, beyond IR, with an overall risky-cardiometabolic profile in children. Serum protein levels of vasorin (VASN), one of the most promising novel biomarkers identified in this study, were further associated with IR in the pubertal stage.

**Conclusions:** To our knowledge, this is the first longitudinal multi-omics approach characterizing molecular blood alterations for IR and obesity during the metabolically critical period of puberty. Our results shed light on the molecular mechanisms underlying epigenetic alterations in obesity and propose novel and promising biomarkers for IR and metabolic alterations in children.

## Background

Insulin resistance (IR) is a pathological condition of glucometabolic sufferance contributing to type 2 diabetes and cardiovascular disease (CVD) in both adults and children. Of note, obesity is the main driver of IR in children [1, 2]. Many children who are overweight or suffer from obesity before puberty maintain obesity in early adulthood, which is associated with increased morbidity and mortality [3–5]. The high mortality rates among people with obesity are mainly due to the development of type 2 diabetes and the increased risk of CVD [6]. Therefore, it is crucial to prevent and treat obesity and IR from the early periods of life [7].

Puberty is a period characterized by dynamic physiological changes, including activation of the reproductive axis and subsequent increase in sex steroids secretion, acceleration in growth, and accumulation of both lean and fat mass [8]. Besides physiological events, puberty has also been associated with differential disease prognosis for conditions such as IR, reinforcing the relevance of this development period to life-long health. Nevertheless, pubertal changes seem not to affect all individuals equally [9–11]. In healthy normal-weight youths, there is a drop in insulin sensitivity in mid-puberty, which recovers at puberty completion. In youth who are obese going into puberty, otherwise, there is evidence that such IR does not resolve, which may result in increased cardio-metabolic risk. Accordingly, youth-onset type 2 diabetes incidence is also tightly linked with pubertal development [12]. Understanding the molecular and biological processes underlying metabolic changes during puberty and the additional impact of obesity on these changes is crucial for preventing type 2 diabetes. Thanks to that, novel non-invasive early diagnostic markers could arise with a great utility for reducing obesity-associated mortality.

DNA methylation (DNAm) is a heritable epigenetic mark consisting of the covalent addition of a methyl group to a cytosine followed by a guanine (CpG). DNAm is potentially reversible and can be altered by environmental factors, resulting in gene expression alterations and providing an interactive connection between genetics, specific diseases and the environment. Indeed, differential DNAm in certain loci has been related to obesity [13], systemic IR [14–22], and type 2 diabetes [13, 15, 16, 23–27] in adults, either in blood or in other metabolically relevant tissues. During puberty, the dynamics of DNAm have also been investigated in one or both genders, emphasizing how DNAm is stable at some CpG sites and varies at others [28, 29]. On the other hand, transcriptional dysregulation of genes has been reported as a key molecular mechanism associated with IR and obesity, possibly connected to DNAm alterations [30, 31]. In this regard, there is evidence of the interacting effects between DNAm and gene expression and the risk for glucometabolic alterations in adult women with obesity (phenomena known as expression quantitative trait methylations (eQTMs)) [17].

Although recent genome-wide association studies (GWAS) have identified numerous single nucleotide polymorphisms (SNPs) associated with type 2 diabetes and its related traits [32–38], these variants only explain a small proportion of the estimated heritability (15–18 %), proposing that there are additional genetic factors left to be discovered. Among the best explanations, it highlights the existence of interacting phenomena between SNPs and DNAm epigenetics marks, known as methylation quantitative trait locis (mQTLs). Interestingly, a previous study demonstrated how interactions between SNPs and DNAm influence mRNA expression and insulin secretion in adult human pancreatic islets [39].

Although the understanding of the molecular and biological processes underlying IR in obesity is growing (especially in adults), none is known about the omics alterations characterizing IR in obesity during the metabolically critical period of puberty, and how it might contribute to the increased disease risk (e.g., for type 2 diabetes). For this purpose, multi-omics approaches present as a promising resource in which systems biology can be applied to mine the complex interactions between genetics, epigenetics, and transcriptomics. The identification of multi-omics signatures and how they relate with the progression of obesity and IR during puberty will allow us to provide pediatricians with robust non-invasive early biomarkers for type 2 diabetes risk. For this task, longitudinal designs are also encouraged.

Considering all this, in the present study, we have identified the multi-omics signatures (DNAm in CpGs, eQTMs and mQTLs) associated with IR in children with obesity, before, during, and after the onset of puberty. This research is a continuation of the PUBMEP study, which evaluates the prevalence of metabolic syndrome and the progression of the cardio metabolic risk factors related to it, from pre-puberty to puberty, in a longitudinal cohort of Spanish children [40].

## Research Design and Methods

### 2.1. Study population

This analysis was conducted within the context of the PUBMEP study. The main clinical findings derived from the PUBMEP study and additional details on the whole study cohort have been already published and are available elsewhere. In the PUBMEP study, all children were first recruited as prepubertal children during 2012–2015 and called again for follow-up medical consultation in 2018. At the moment of recruitment, children were aged 4–12.1 years and came from three Spanish recruiting centers (cities). At the second visit, children were aged 9.72–18.07. All subjects with clinical signs of reached puberty were finally included in the longitudinal population. During the course of the study (2012–2018), children remained under regular medical monitoring by the same pediatricians. The assessment of the pubertal stage was carried out following the Tanner classification [41] and confirmed with a hormonal study. Here, a sub-population of 139 children (76 females) from the whole PUBMEP cohort was selected for omics analyses. The main inclusion criterion for this sub-project was presenting a good-quality DNA sample in the prepubertal stage. The following characteristics were considered as exclusion criteria: birth weight <2500 g; intake of any drug that could alter blood glucose, blood pressure or lipid metabolism; not being able to comply with the study procedures and being participating or having participated in the last three months in an investigation project. The 139 participating children were organized in a longitudinal approach of 90 children (47 females) and two cross-sectional approaches of 99 (52 females) and 130 (71 females) children for prepubertal and pubertal stages, respectively. A general overview of the study design, populations and statistical analyses performed is well-described in **Figure 1**. The longitudinal approach consisted of 90 Spanish children (47 females) allocated into five experimental groups according to their obesity and IR status before and after the onset of puberty (**Figure 2A**).

**Figure 1.**
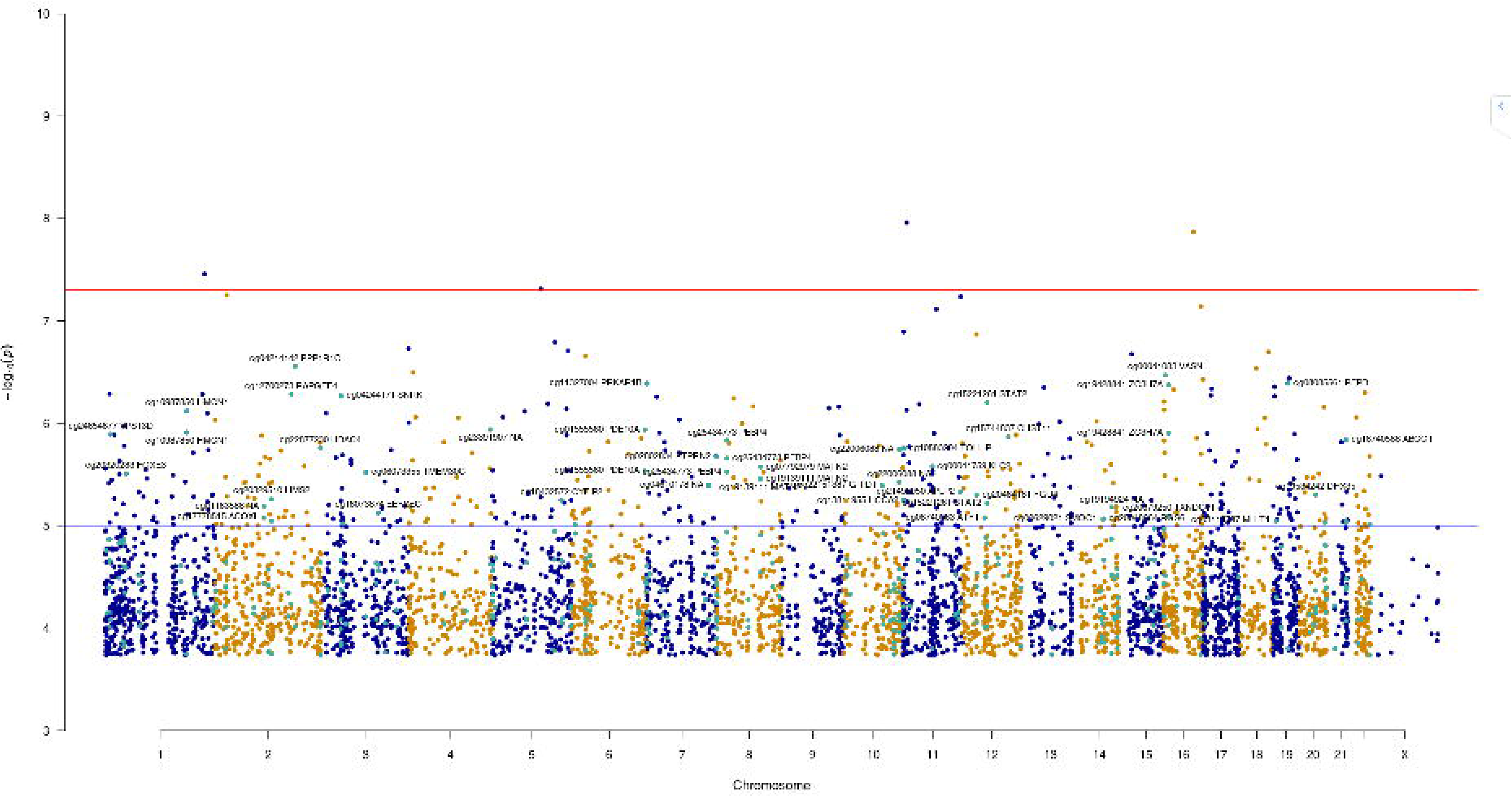
Overview of the study design, populations under study and statistical analyses performed.

**Figure 2.**
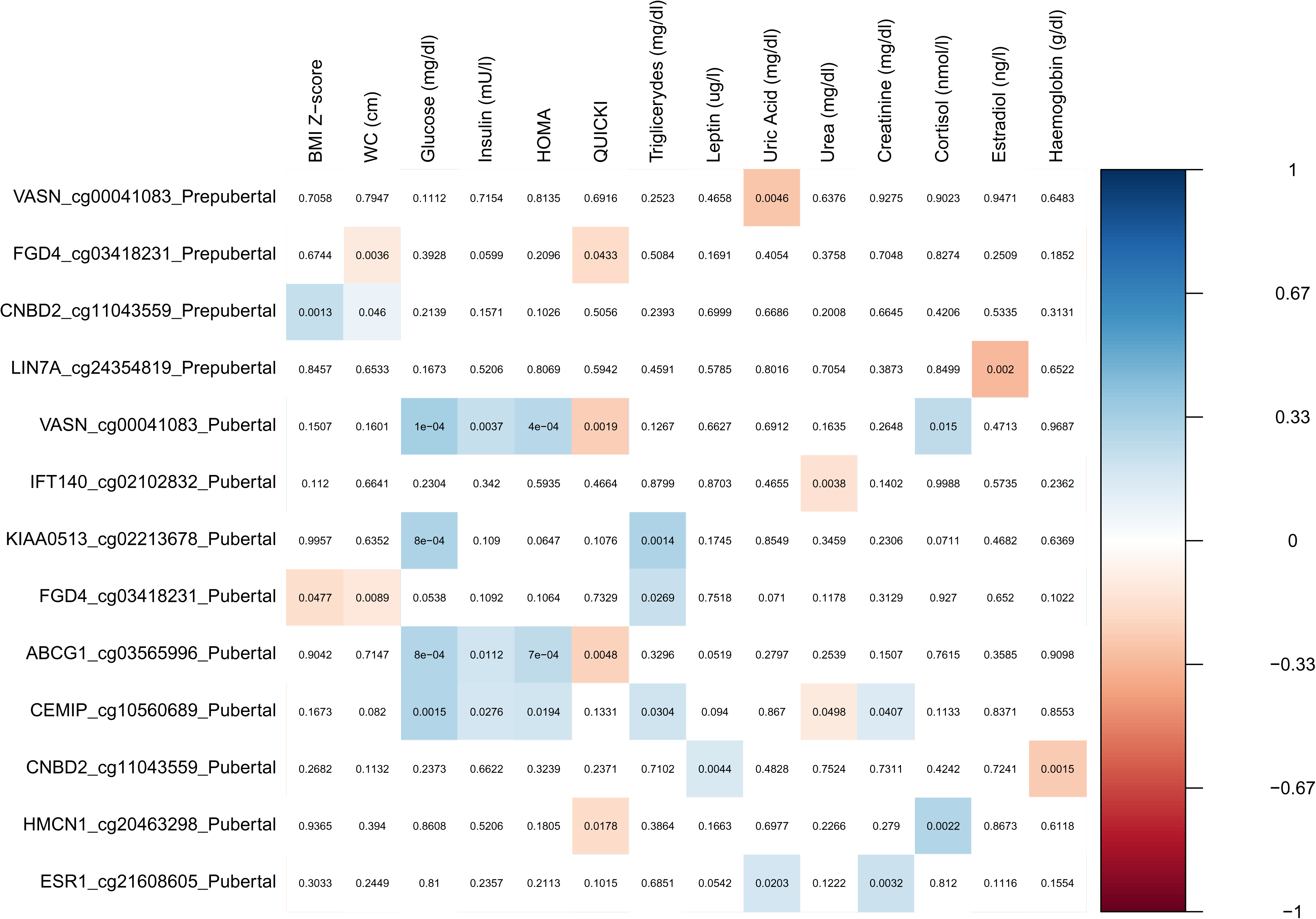
The statistical design adopted for DNA methylation bioinformatics analysis of IR.

All participants (N=139) presented DNA samples with enough quality for genomics (GWAS) and epigenomics (EWAS) analyses. Moreover, we also collected blood samples in 44 children of the pubertal stage using PAXGEN-RNA tubes for posterior RNA-seq analysis. Descriptive statistics for these longitudinal and cross-sectional approaches are available in the **Additional files 1, 2 and 3**. The main blocks of analyses performed in this work consisted of 1) EWAS analysis of IR and 2) multi-omics integration of EWAS results along with GWAS and RNAseq.

### 2.2. Ethics statement

These studies were conducted following the Declaration of Helsinki (Edinburgh 2000 revised), and they followed the recommendations of the Good Clinical Practice of the CEE (Document 111/3976/88 July 1990) and the legal in-forced Spanish regulation, which regulates the clinical investigation of human beings (RD 223/04 about clinical trials). Accordingly, the corresponding ethics committees approved the study at each of the participating centers (Code IDs GENOBOX: Córdoba01/2017, Santiago 2011/198, Zaragoza 12/2010; and PUBMEP: Córdoba 260/3408, Santiago 2016/522, Zaragoza 22/2016, Granada 01/2017).

### 2.3. Anthropometry, biochemical measurements, and inflammation and cardiovascular risk biomarkers

Anthropometric measurements such as body weight (kg), height (cm), hip circumference (cm) and waist circumference (WC) (cm) were measured at each time point using standardized procedures, and BMI (kg/m^2^) was calculated. BMI z-score was estimated based on the Spanish reference standards published by Sobradillo et al. [42]. Blood pressure was measured three times for each individual by the same examiner using a mercury sphygmomanometer and following international recommendations [43]. Measures of lipid and glucose metabolism, hormones and classical biochemical parameters were performed at the laboratories of each participating hospital following internationally accepted quality control protocols.

Blood samples from both time points were collected in overnight fasting conditions, centrifuged, and plasma and serum were stored at-80°C. Plasma adipokines, inflammation, and cardiovascular risk biomarkers (adiponectin, leptin, resistin, tumor necrosis factor alpha (TNF-α), high-sensitivity CRP (hsCRP), interleukin (IL)-6, IL-8, total plasminogen activator inhibitor-1 (PAI-1), P-Selectin, myeloperoxidase (MPO), monocyte chemoattractant protein 1 (MCP-1), matrix metalloproteinase-9 (MMP-9), soluble intercellular cell adhesion molecule-1 sICAM-1, and soluble vascular cell adhesion molecule-1 (sVCAM)) were analyzed in all samples and time points using XMap technology (Luminex Corporation, Austin, TX) and human monoclonal antibodies (Milliplex Map Kit; Millipore, Billerica, MA) as previously reported [44, 45]. S100A4 protein levels were determined in plasma using the CSBEL02032HU (Cusabio Biotech Co, Ltd., Wuhan, China), an enzyme-linked immune-absorbent assay kit according to the manufacturers’ instructions. The coefficient of variance was 7%.

### 2.4. HOMA-IR cut off points

The IR status was here defined by means of the HOMA-IR index. Since HOMA-IR strongly varies between ages, genders and diseases, and since no reference values have been yet established in either children or adult populations [46, 47], own cut-off points were extracted from a previous well-described Spanish cohort composed of 1669 children and adolescents [44, 48]. For the prepubertal stage, a single cut-off value of HOMA-IR=2.5 was considered for IR [44, 48]. For the pubertal stage instead, gender information was taken into consideration and different cut-off points were adopted for IR according to the 95^th^ HOMA-IR percentile. Extracted from a subset of 778 pubertal Spanish children, cut-off values corresponded to HOMA-IR=3.38 in boys and HOMA-IR=3.905 in girls. These cut-off points have already been tested and validated as good metabolic risk classifiers in our population according to the results from a previous PUBMEP paper [40].

### 2.5. Epigenome-wide association study (EWAS)

EWAS analysis was performed in all children (N=139) and time points, including longitudinal and cross-sectional approaches (**Figure 2**). Buffy coat fractions from blood samples in all children and time points were selected for DNA methylation analysis. Genomic DNA was extracted from peripheral white blood cells using two automated kits, the Qiamp DNA Investigator Kit for coagulated samples and the Qiamp DNA Mini & Blood Mini Kit for non-coagulated samples (QIAgen Systems, Inc., Valencia, CA, USA). All extractions were purified using the DNA Clean and Concentrator kit from Zymo Research (Zymo Research, Irvine, CA, USA). High-quality DNA samples (≥ 500 ng) were treated with bisulfite using the EZ-96 DNA Methylation Kit (Zymo Research Corporation, Irvine, CA). DNA methylation was measured with the Infinium Methylation EPIC array using bead chip technology (Illumina, San Diego, CA, USA).

Raw intensity signals from IDAT files were loaded into the R environment using the *MINFI* R package. As a result, we obtained an RGChannelSet object containing all the raw intensity data, from both the red and green color channels, for each of the samples and time records. We generated a detection p-value for every CpG in every sample by comparing the total signal for each probe to the background signal level, which was estimated from the negative control probes. To minimize the unwanted variation within and between samples, we applied Beta-Mixture Quantile (BMIQ) intra-array normalization, including all individuals and time records. Poor performing probes were filtered out according to different criteria: probes with a detection p-value above 0.01 in more than 10 % of the samples (number of probes= 230), probes with SNPs (number of probes= 30,432), cross-reactive probes aligning to multiple locations (number of probes= 25,570) and probes located on the Y chromosome (number of probes= 246). After applying all these filters, 809,381 probes remained in the dataset.

As methylation is cell type-specific and methylation arrays provide CpG methylation values for a population of cells, biological findings from samples comprised of a mixture of cell types, such as the case of peripheral blood, can be confounded due to variable cell-type composition. To correct analyses for the variable proportion of each white cell type in our subjects, we employed the reference EPIC 850k dataset published by Salas et al. (2012) and the Houseman procedure [49]. The influence of each confounding variable on the global state of methylation in our population was assessed by means of correlation studies and heatmap plots using the *SWAMP* R package v1.4.1.

Intensity values were used to determine the proportion of methylation at each CpG site. Methylation levels were reported as either beta values (β-Values = M/(M + U)) or M-values (M value = log2(M/U)), where M and U correspond to the Methylated and Unmethylated signals, respectively. Beta values and M-values are related through a logit transformation (M-value = log2(β-value /1-β-value)). Because percentage methylation is easily interpretable, beta values in the present paper were employed for describing the level of methylation at each locus and for graphical presentation of results. On the other hand, due to their distributional properties, M-values were selected for statistical testing. All described analyses were performed in R environment version 4.0.3.

### 2.6. Genome-wide association study (GWAS)

GWAS analysis was performed in all children (N=139) (**Figure 1**). When two samples of DNA were available for the same individual (e.g., in the longitudinal approach), the most recently extracted DNA sample was selected for genomics analysis. Whole-genome genotyping analysis was performed on the i-SCan platform using the Infinium HTS Assay (Illumina, San Diego, CA, USA). The Bead Chip selected for the project was the Infinium Global Screening Array-24 v3.0 Kit, which includes ∼ 654,000 genetic markers associated with complex diseases. After quantification of DNA samples by fluorimetry, they were normalized to 200-400 ng of DNA per sample in deep well plates, as established in the Infinium HTS Assay Protocol.

The first step of the primary data analysis consisted of the extraction of genotype calls from fluorescence data and the construction of work data files for data manipulation and analysis. Using the *GenomeStudio* software, we obtained genotype calls for all individuals and generated the standard format files (*.ped* and *.map*). Data were then imported into *PLINK* 1.9 software [50], and converted into binary format files using the *--make-bed* flag. These binary formats *(.bed*, *.bim* and *.fam*) are a more compact representation of the data that saves space and speeds up subsequent analyses. We implemented a quality control (QC) process in PLINK 1.9 software before high-level statistical analyses. According to literature, we applied standard QC filters including: 1) Exclusion of SNPs and individuals with a missing data rate >= 10%, and 2) Exclusion of SNPs with a minor allele frequency (MAF) < 1% or a Hardy-Weinberg Equilibrium (HWE) P-value < 1×10^−5^ in controls. As a result, 471,192 SNPs remained in the dataset. These filters were selected in accordance with procedures described elsewhere [51] to minimize the influence of genotype-calling artefacts in a GWAS.

### 2.7. Next-generation transcriptome sequencing (RNA-Seq)

RNA-seq analysis was performed in a subset of 44 children from the pubertal cross-sectional approach (**Figure 1**), which had also been included in the EWAS and GWAS. RNA was extracted from peripheral blood using the PAXgene® Blood RNA Kit (PreAnalytiX/QIACUBE) according to the manufacturer’s instructions. The concentration and quality of extracted RNA were measured using the Qubit 4 Fluorometer (Thermo Fisher Scientific, MA, USA) and the 2100 Bioanalyzer Instrument (Agilent Technologies, CA, USA). Libraries from mRNA were prepared g of RNA starting material and the TruSeq Stranded mRNA Library Prep Kit μ (Illumina, CA, USA) according to the manufacturer’s protocol. This protocol captures poly-adenylated RNA by transcription by oligo-dT primer, after which the RNA is fragmented. The sample is back transcribed to generate the cDNA, both in the first and second strands. The 3’ ends are adenylated, the adapters and barcodes are ligated, and finally, the sample is enriched by PCR. Adapters and sample codes (index-barcodes) are added to the libraries to be simultaneously sequenced. mRNA libraries were sequenced on the Next-Seq 500 system (Illumina, CA, USA) using the highest output mode and paired-end 75 bp read lengths with a depth of 20 million reads for each sample. To get a depth of 20 million reads per sample 2 runs with 4 lanes for each run were conducted.

Primary RNA-seq bioinformatics analyses were implemented in R environment separately for each run following standardized published recommendations. Primary analyses included processing raw sequencing reads, aligning to the reference genome, and quantitating the expression levels. The aligning of RNA-seq reads to the genome was conducted using *HISAT* software 2.2.1 release [52]. We sorted and converted the generated SAM files into BAM using *SAMTOOLS* [53]. Then, we used the *FEATURECOUNTS* R package [54] to generate count matrices from reads aligned to the genome. As reference genome, we used the *hg38* version from the Ensembl [55]. From it, we created a transcript database, using the function *makeTxDbFromGFF* from the *GENOMICFEATURES* R package. Finally, we obtained two datasets of 60,058 quantified ENSG ids (one per run). After confirming the grouping of technical replicates (samples) among runs by PCA plot (Additional file 4), we merged the two counts datasets by applying a sum.

### 2.8. Descriptive statistics

At each cross-sectional approach (**Figure 2B**), continuous non-omics variables were tested for normality using the Shapiro–Wilk test. Heteroscedasticity between experimental groups was explored through the Levene test. T-tests and Mann–Whitney U-tests were applied conveniently to determine group differences at each cross-sectional stage (prepubertal and pubertal). The resulting descriptive statistics are available in **Additional files 2 and 3**. In the longitudinal approach (**Figure 2A**), within-group changes from prepuberty (T_0_) to puberty (T_1_) were assessed using a paired design, employing either a paired t-test or a Wilcoxon signed-rank test. Between-group differences were assessed by one-way ANOVA, Kruskal-Wallis or Welch tests to the computed delta values (T_1_–T_0_) for each continuous measurement according to standard statistical assumptions. The one-way ANOVA, Kruskal-Wallis and the Welch test were also employed to assess group differences (among the 5 experimental groups) at each stage (time point) of the longitudinal approach. Pairwise t-tests, pairwise Mann– Whitney U-tests and Dunn tests were applied conveniently as post-hoc analyses to determine which experimental groups differed from each other in these analyses. The resulting descriptive statistics are available in **Additional file 1**. All described analyses were performed in R environment version 4.0.3 [56].

### 2.9. DNA methylation bioinformatics analysis on insulin resistance

After pre-processing, 809,381 CpGs probes passing quality filters in the EWAS were selected for high-level statistical analyses. The statistical design adopted for DNA methylation bioinformatics analysis of IR is presented in **Figure 2**. The main objective of this analysis was identifying the DNA methylation patterns associated with IR development, amelioration or worsening in children with obesity during the onset of puberty. For avoiding confounding with non-pathological aging epigenetics marks, derived findings were contrasted to the DNA methylation patterns associated with the onset of puberty in the normal-weight group *G1*. For the longitudinal approach (N=90), we investigated the changes in DNA methylation from prepuberty to puberty of each included CpG site performing both within-and between-groups comparisons. Within-group changes in DNA methylation from prepuberty (T_0_) to puberty (T_1_) were studied exclusively for the groups *G3* and *G4*, since these were the only groups presenting changing trajectories for IR. Between-groups changes in DNA methylation from prepuberty (T_0_) to puberty (T_1_) were otherwise investigated for all pairwise group combinations that involved either the *G3* or *G4* group (for more details see **Figure 2A**). These analyses were implemented in the R environment using linear models from the *LIMMA* R package and considering the M-values of DNA methylation for each CpG as the outcome or dependent variable. For that purpose, a multi-level experiment was considered, treating the patient as a random effect, and the experimental group and time as a combined fixed factor. The inter-subject correlation was the input for the linear model fit. Contrasts of interest over the constructed linear model were then applied using a moderated t-test. All this was implemented in *LIMMA* using the functions *duplicateCorrelation*, *lmFit*, *contrasts.fit* and *eBayes*. Analyses were conveniently adjusted for confounders, including gender, city of origin, blood cell proportions and age. Raw p-values were corrected using the false discovery rate (FDR) according to the Benjamin-Hochberg procedure for multiple comparisons.

In cross-sectional approaches (N=99 and N=130) (**Figure 2B**), the objective was also focused in identifying the DNA methylation patterns associated with IR, but including now those marks associated with IR before the onset of puberty. Since cross-sectional approaches included extra children with regard to the longitudinal cohort of 90 children (**Figure 1**), these analyses were also intended as validation approaches for the longitudinal findings. In these analyses, *LIMMA* linear models were conducted with DNA methylation levels as outcome, experimental group as a categorical predictor and gender, city of origin, blood cells proportions, batch, obesity status (when necessary) and age as covariates. Raw p-values were corrected using the false discovery rate (FDR) according to the Benjamin-Hochberg procedure for multiple comparisons.

We selected exclusively overlapping findings from these analyses and different approaches (**Figure 3**), obtaining a curated list of loci whose methylation is robustly and repeatedly associated with IR in our design. This list of IR-associated CpGs (and their mapped genes) (thereinafter known as ‘validation-list’) was the input for the next blocks of analysis. For this selection, we first created a Venn diagram including the genes reported as significant in each statistical approach. Once overlapping IR-associated genes were identified, all significant CpGs mapping these genes were selected. The resulting list was composed of 267 IR-associated CpGs mapping 128 genes.

**Figure 3.**
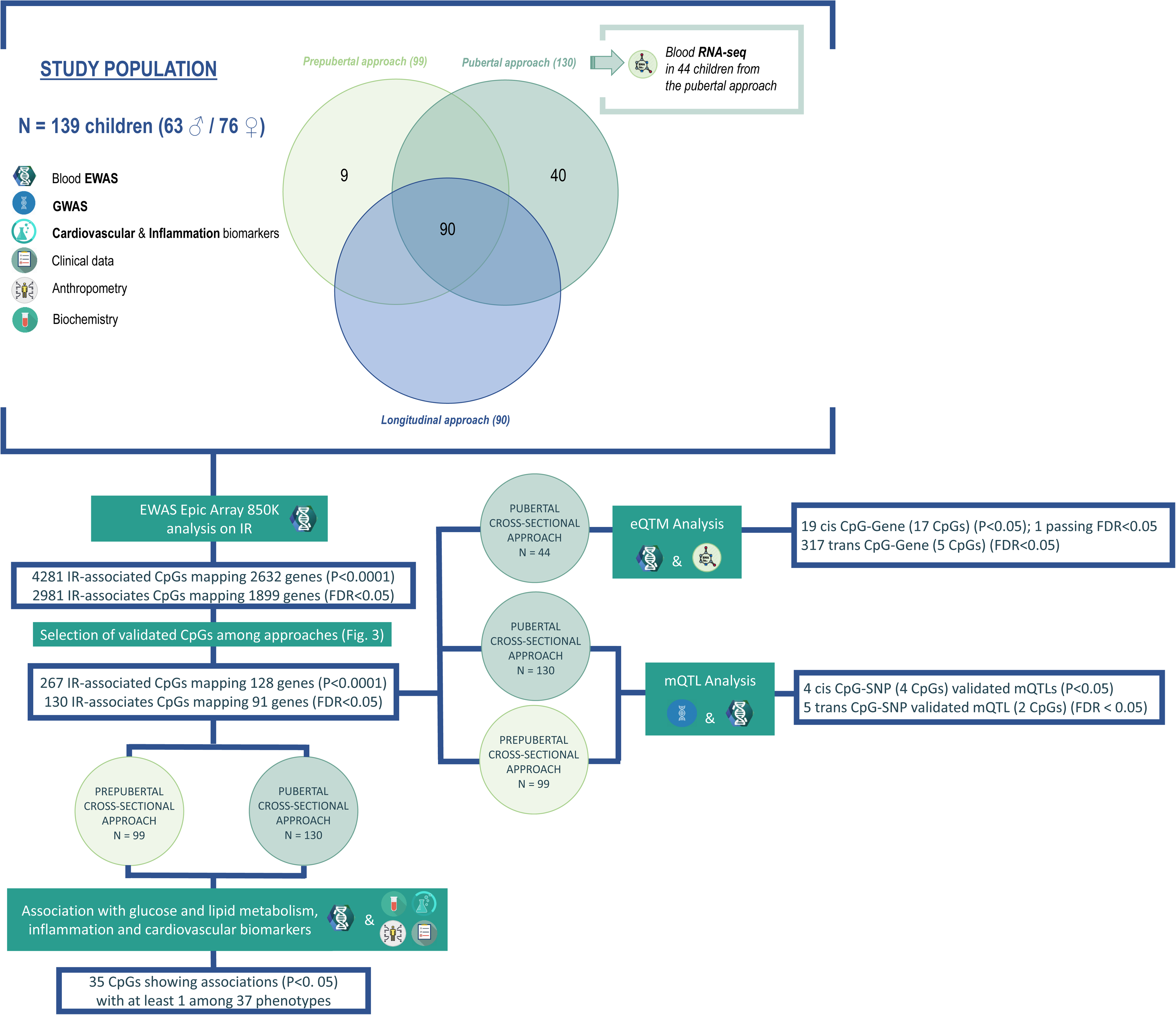
Overlapping findings (at loci level) between the statistical approaches of the EWAS on insulin resistance. These findings constitute the so called ‘validation-list’.

In cross-sectional approaches, we further applied multiple linear regressions (MLR) between DNA methylation M-values of the CpGs in the ‘validation-list’ and continuous non-omics outcomes (including all collected anthropometric and biochemical measurements, and inflammation and cardiovascular biomarkers) at each temporal record (T_0_ and T_1_). The main purpose of these additional analyses was to identify those CpGs, which instead or besides IR, are associated with other obesity-related metabolic alterations or parameters. In these analyses, IR, BMI Z-score, gender, age, city of origin and height were included as covariates conveniently. Given the number of analyzed outcomes, we again considered FDR as in Benjamin-Hochberg to correct for multiple hypothesis testing.

### 2.10. Expression quantitative trait methylation (eQTM) analyses

The expression quantitative trait methylation (eQTM) analysis is intended for identifying gene expression regulatory phenomena, in which the methylation of a CpG influence (up-or down-regulating) the expression of a target transcript. For these analyses, we used linear regression, as implemented in the *MatrixEQTL* R package, to test whether DNAm levels of the CpG sites from the ‘validation-list’ are associated with transcript expression levels (considering all the transcripts mapped by our RNA-seq analysis). For these analyses, gene expression data for 60,058 transcripts were normalized using the quantile normalization method. Here, the transcript expression level (normalized) was the outcome and the methylation level (M-value) of each CpG site was the predictor, with gender, age and city of origin as adjusting covariates. We searched for cis-eQTMs in and around 10,000 bp of each transcript and trans-eQTMs if the distance between the CpG and the transcript was higher than 10,000 bp. As a measure of eQTMs effect size, we reported the beta regressors estimated by the linear model. The p-values from the linear regression analysis were adjusted for multiple comparisons using the Benjamini-Hochberg FDR procedure. These analyses focused on 267 CpGs and 60,058 high-quality transcripts as input. In the cis-analysis, no correction value was used for correcting multiple testing. The correction value for the trans-analysis was calculated as the total number of analyzed CpG sites multiplied by the number of transcripts in the whole dataset.

### 2.11. Methylation quantitative trait loci (mQTL) analysis

The methylation quantitative trait loci (mQTL) analysis is intended for identifying epigenetics regulating phenomena, in which an SNP regulates the methylation levels of a CpG. These phenomena could be, therefore the molecular explanation for some epigenetics IR-associated marks for which the environment is not the causal mediator. For these analyses, we used linear regression, as implemented in *MatrixEQTL* R package. In the linear model, DNA methylation values were modelled as the outcome, SNP genotypes from GWAS were encoded as 0, 1 or 2 according to the number of minor alleles (additive genetic model), and gender, age and city of origin were included as covariates. To distinguish between local (cis-mQTLs) and distant (trans-mQTLs), an arbitrary boundary with the maximum distance of 500 bp between SNPs and CpG sites was used to define cis-mQTLs. All other SNP-CpG pairs were considered as trans-mQTLs. P-values were adjusted with a correction value for multiple testing, which considers the dependency of linkage disequilibrium (LD) between SNPs by LD-based pruning and thereby uses the number of independent tests. In the cis-analysis, no correction value was used for correcting multiple testing. The correction value for the trans-analysis was calculated as the total number of analysed CpG sites multiplied by the number of SNPs in the whole dataset.

### 2.13. Reference genome assembly

EWAS, GWAS and RNA-seq datasets were functionally annotated based on GO and KEGG ontologies using entrez gene identifiers and the database *org.Hs.eg.db* [57]. For eQTM and mQTL analyses, the three employed omics datasets were re-annotated or flipped (in terms of chromosome number and genomic locations) to the same assembly (*hg38*) as reference.

### 2.14. Functional annotation of CpG sites and regions

Selected CpG sites and regions were further annotated using the *ILLUMINAHUMANMETHYLATIONEPICANNO.ILM10B4.HG19* and *MISSMETHYL* R packages. Genes associated with each CpG site were obtained using the *getMappedEntrezIDs* function. The annotation in terms of genomics regulatory elements consisted of two categories: 1) distance to a CpG island and 2) annotation to gene region. The distance related annotations identify whether CpG sites overlap a known CpG island, 2000bp of the flanking regions of the CpG islands (shores), 2000bp of the flanking regions of the shores (shelves), or outside these regions (open sea). CpGs overlapping gene bodies were annotated as (Body). The gene region analysis classified CpGs in the context of genes, namely, exons, UTRs, introns, promoters, and intergenic regions. Additional annotation of CpG sites for nearby SNPs was determined using the UCSC database. To identify the regulatory potential of CpG sites, each site was categorized based on its predicted chromatin state. These data and additional information have been gathered for significant CpGs and are available in each table result.

### 2.15. Gene ontology and biological pathway enrichment analysis

The *gometh* function from the R package *MANIFEST* was used to determine enrichment of CpG-annotated genes in KEGG terms, biological pathways, and cellular and molecular functions. This function takes a character vector of significant CpG sites, maps the CpG sites to Entrez Gene IDs, and tests for GO term or KEGG pathway enrichment using a hypergeometric test [58], taking into account the number of CpG sites per gene on the EPIC array. The *gometh* is based on the *goseq* method [59] and calls the *goana* function from the *LIMMA* R package [60]. The *gometh* tests all GO or KEGG terms, and FDR are calculated using the method of Benjamini and Hochberg (1995). The *LIMMA* functions *topGO* and *topKEGG* were used to display the top most enriched pathways.

## Results

### 1. Clinical characteristics of participants

A general overview of the study design, populations and statistical analyses conducted is well-described in **Figure 1** and **Figure 2**. Descriptive statistics for longitudinal and cross-sectional approaches are available in the **Additional files 1, 2 and 3**. **Figure 2A** describes all details on the longitudinal approach. At baseline, there was no age difference between experimental groups, with all but the G5, which maintains IR with pubertal maturation, presenting not significant differences in the elapsed time between visits. As expected from the study design, groups maintaining or developing IR with the onset of puberty, G4 and G5, presented the highest increases in HOMA-IR and fasting serum insulin as compared with the insulin-sensitive groups (**Additional file 1**). Among them, the G4 group, which develops IR with the onset of puberty, also displayed the highest increase in waist circumference, hip circumference and SBP, and inflammatory and cardiovascular biomarkers (e.g., P-selectin and t-PAI) (**Additional file 1**). Interestingly, the G3 group, presenting the opposite behavior for IR (amelioration) than G4, showed a decrease in plasma glucose, triacylglycerol concentrations and MCP1 cytokine levels, and these changes were significantly different from the patterns observed in the rest of the groups. Thus, the experimental groups of our design were representative of the longitudinal IR and insulin-sensitive trajectories during the onset of puberty (**Additional file 1**). As expected from the experimental design, an overall decrease in total and low-density lipoprotein cholesterol levels during puberty was observed for all groups, as well as a decrease in adiponectin concentrations. Likewise, cross-sectional experimental groups showed coherent behaviors in anthropometry, glucose, lipid, inflammatory, and cardiovascular biomarkers (**Additional files 2 and 3**).

### 2. Blood cells CpG methylation is associated with obesity insulin resistance

After pre-processing, 809,381 CpGs EWAS probes passing quality filters were selected for high-level statistical analyses. The statistical design adopted for DNA methylation analysis of IR is presented in **Figure 2**. The main objective of this design was to identify the DNA methylation patterns associated with IR development, amelioration or worsening in children with obesity before, during, and after the onset of puberty. Our analysis identified 4,281 IR-associated unique CpG sites (P-value < 0.0001), from which 2,981 further presented an FDR < 0.05 (**Additional file 5**). Annotation of differentially methylated sites (DMS) informed that reported CpGs were linked to 2,632 and 1,899 genes, respectively for raw P-value and FDR thresholds. We assessed the robustness of our findings by comparing our list of 2,632 IR-associated genes to a curated list of genes whose methylation degree is strongly associated with type 2 diabetes according to a recent large EWAS meta-analysis conducted in Europeans adults [23]. Our list contained 8 of the 38 type 2 diabetes-associated EWAS genes validated by *Juvinao et al*. 2021 [23] (*ABCG1*, *CDH23*, *CPT1A*, *HCCA2*, *HDAC4*, *KRT4*, *PBX1* and *SGK2*), highlighting three of them among the top 6 associated genes in the meta-analysis. Other 24 well-known diabetes and obesity epigenetics loci recently reviewed by *Ling C et al.* 2019 [13], were also present among our associations (*ABCC3, ADCY5, ATP10A, CDKN1A, CXCL14, DNMT3A, FADS2, FTO, GLP1R, GRB10, HIF3A, KCNQ1, MALT1, MOGAT1, NCOR2, NFAM1, PLCB1, PPARG, PRDM16, PRKCE, SEPT9, TCF7L2, THADA* and *VAC14*). Genes encoding proteins with a key role in the IR-puberty axis and strongly related to the growth hormone, such is the *IGF-1*, were also highlighted in our analysis [61]. Among associations, there were also loci whose DNA methylation, beyond type 2 diabetes, have been specifically associated with IR according to literature (*COL18A1, CTNND2, CXCL1, DNMT3A, GRB10, HDAC4, LAT, PAX6, SH3RF3* and *SIRT2*).

KEGG-pathways enrichment analysis showed that the 2,632 IR-associated genes (passing the raw P-value threshold) were over-represented for pathways with relevance in inflammation and human metabolism in the context of our research including; ‘Ovarian steroidogenesis’, ‘Cortisol synthesis and secretion’, ‘Extracellular Matrix-receptor interaction’, ‘Glucagon signalling pathway’, ‘cAMP signalling pathway’, ‘Insulin secretion’, ‘Phospholipase D signalling pathway’ or ‘PPAR signalling pathway’ (P-value < 0.05) (**Additional file 6**).

As expected from the study design, our analysis also revealed genes previously described as markers of dynamic DNA methylation changes during the course of puberty (e.g., *ADCY9, ATK3, GRIK5, GNG7, PDE10A*, or *TRAF3IP2*) [28].

From the initial list of 2,632 IR-associated genes, we exclusively selected those overlapping findings among statistical approaches (**Figure 3**), obtaining a reduced list of genes whose methylation is robustly and repeatedly associated with IR in our design. The resulting list of IR-associated genes and their mapping CpGs (thereinafter known as ‘validation-list’) was the input for the next blocks of analysis. This list was composed of 267 IR-associated unique CpGs mapping 128 genes (P-value < 0.0001), from which 130 IR-associated CpGs, mapping 91 genes, presented an FDR < 0.05 (**Figure 4 and Additional file 7**). Association results for the top 25 CpGs from the list are shown in **table 1**. Interestingly, functional enrichment analyses on KEGG terms for these CpGs, conserved interesting biological pathways such as ‘Ovarian steroidogenesis’, ‘cAMP signalling pathway’, ‘Insulin secretion’ and ‘Phospholipase D signaling pathway’, or reported new ones as ‘estrogen signaling pathway’ (P-value < 0.1) (**Additional file 8**). This list also maintained top adult type 2 diabetes loci previously described in the literature (*ABCG1*, *ADCY5*, *DNMT3*, *HDAC4*, *TCF7L2* and *HCCA2*) (RefsLingyJuvinao), and revealed new promising loci never reported as epigenetic marks of IR (e.g. *CDC42BPB*, *ESR1*, *HMCN1*, *PRKAR1B*, *SNRK* and *VASN*, among others). GO-terms enrichment analysis further revealed genes from the ‘validation-list’ mapping important pathways such is the ‘G protein-coupled receptor signalling pathway’ (P-value = 0.002) (**Additional file 9**).

**Figure 4.**
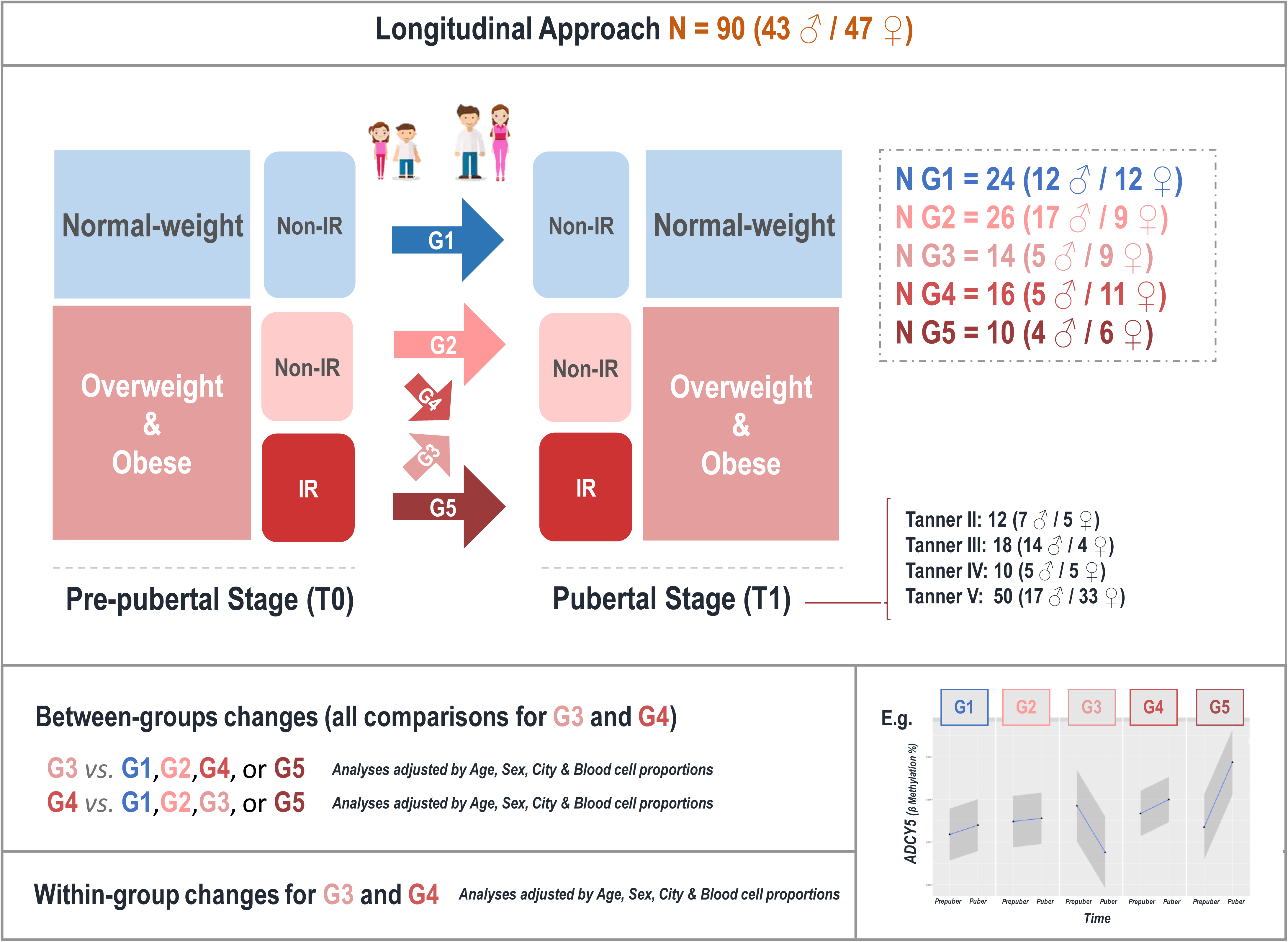
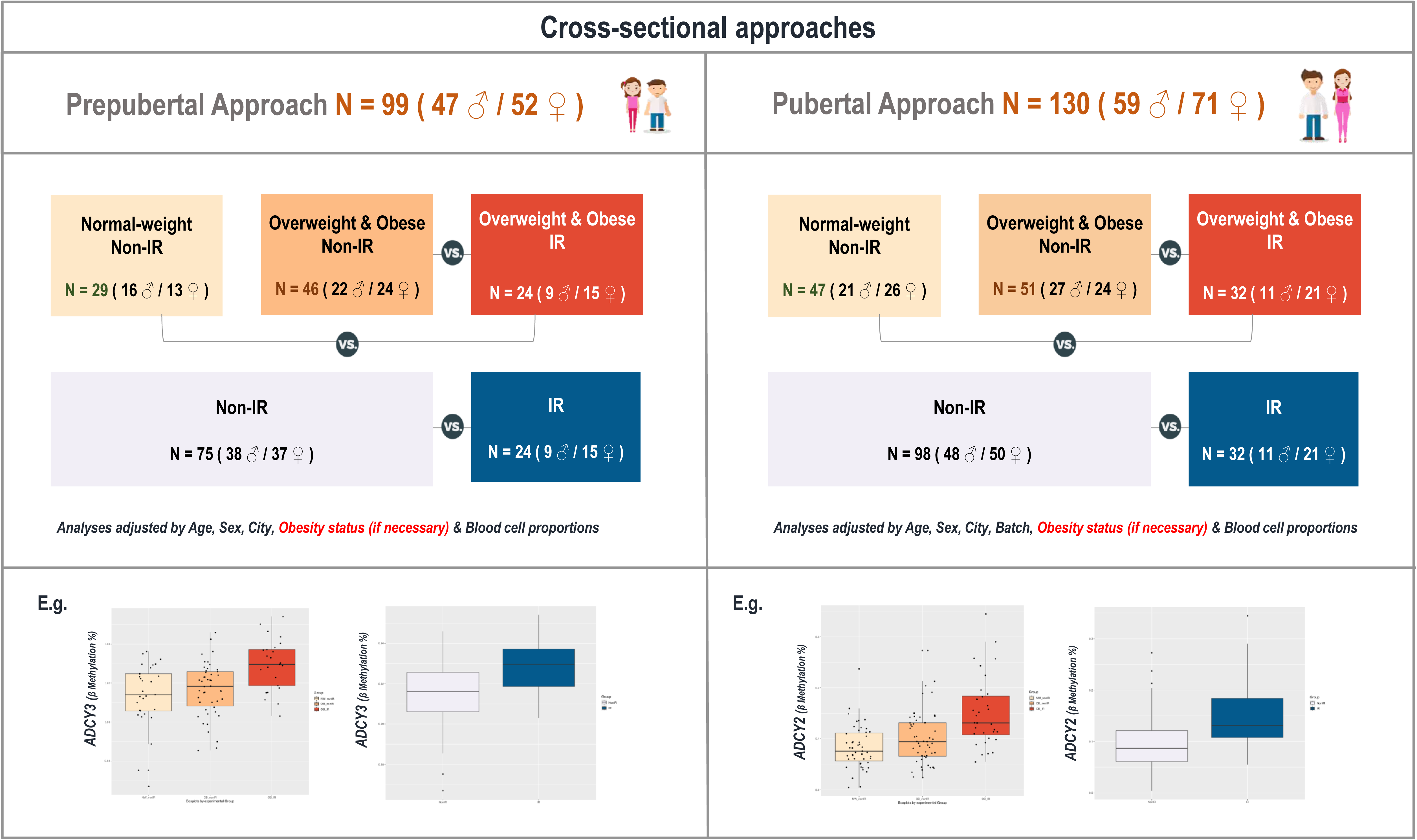
Manhattan plot showing the full list of associations derived from the EWAS on insulin resistance. Loci from the ‘validation-list’ are highlighted in green and labelled.

**Table 1.**
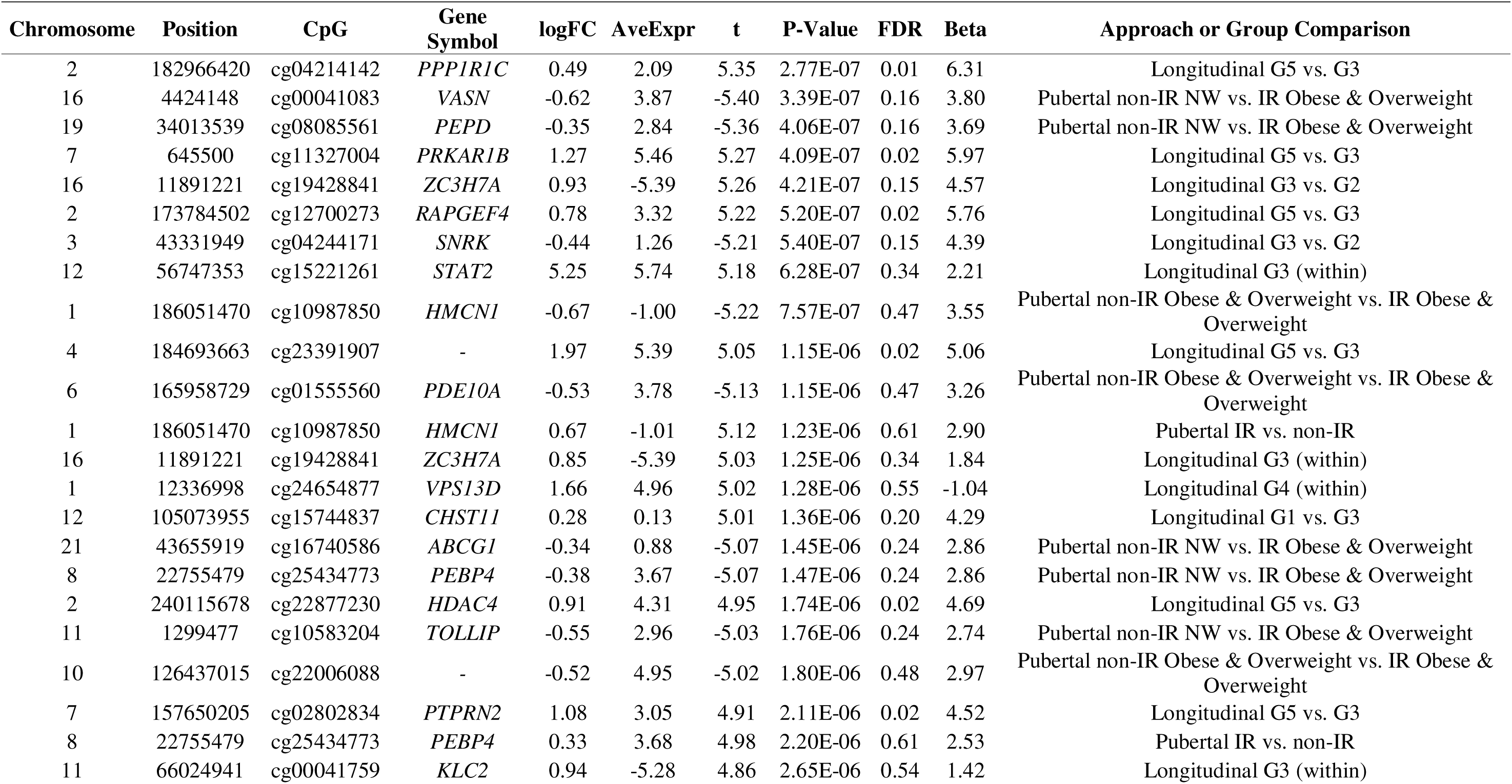
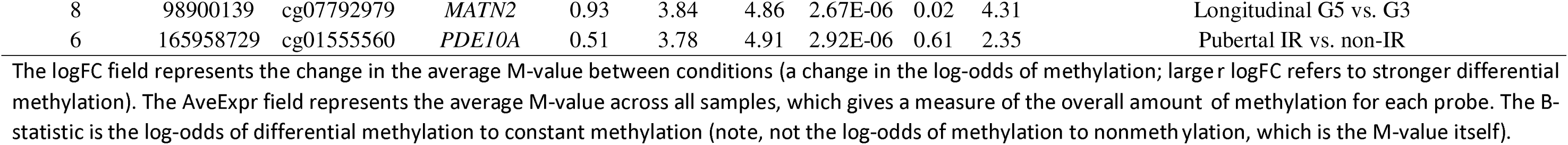
Association results for the top 25 CpGs from the ‘validation-list’. Legend: The logFC field represents the change in the average M-value between conditions (a change in the log-odds of methylation; larger logFC refers to stronger differential methylation). The AveExpr field represents the average M-value across all samples, which gives a measure of the overall amount of methylation for each probe. The B-statistic is the log-odds of differential methylation to constant methylation (note, not the log-odds of methylation to nonmethylation, which is the M-value itself).

### 3. Association between DNAm and other phenotypic traits at key CpGs identified by our EWAS

At each cross-sectional approach, we applied multiple linear regressions to the CpGs from the ‘validation-list’ and continuous non-omics outcomes (including all collected anthropometric and biochemical measurements, and inflammation and cardiovascular biomarkers). The main purpose of these additional analyses was to identify those CpGs, which instead or besides IR, are associated with other obesity-related metabolic alterations or parameters. In **additional file 10**, we present all significant associations showing a P-value < 0.05 with at least one trait after confounding adjustment. A sub-selection of the top significant associations from these analyses is available in **figure 5** (P-value threshold < 0.005). Interestingly, the methylation degree of three genes showed significant associations with assessed phenotypic traits at both cross-sectional stages (prepubertal and pubertal) (*CNBD2*, *FGD4*, and *VASN)*. Among them, the loci *CNBD2* and *FGD4* reported an association with anthropometry (BMI Z-Score and WC). At the same time, *VASN* reinforced its association with glucose metabolism (glucose, insulin and HOMA-IR) in the pubertal stage. *CNBD2* and *FGD4* also presented associations with metabolic traits in the pubertal stage including, leptin concentrations and QUICKI, and triacylglycerol levels, respectively. Previously literature described genes such as *ABCG1*, or other novels such as *VASN*, *CEMIP* and *HMCN1* reported strongly significant associations with glucose metabolism at the pubertal stage (glucose, insulin and HOMA-IR) in this study. On the other hand, DNAm in the *ESR1* and *VASN* genes was associated with kidney-function markers (creatinine and uric acid levels).

**Figure 5.**
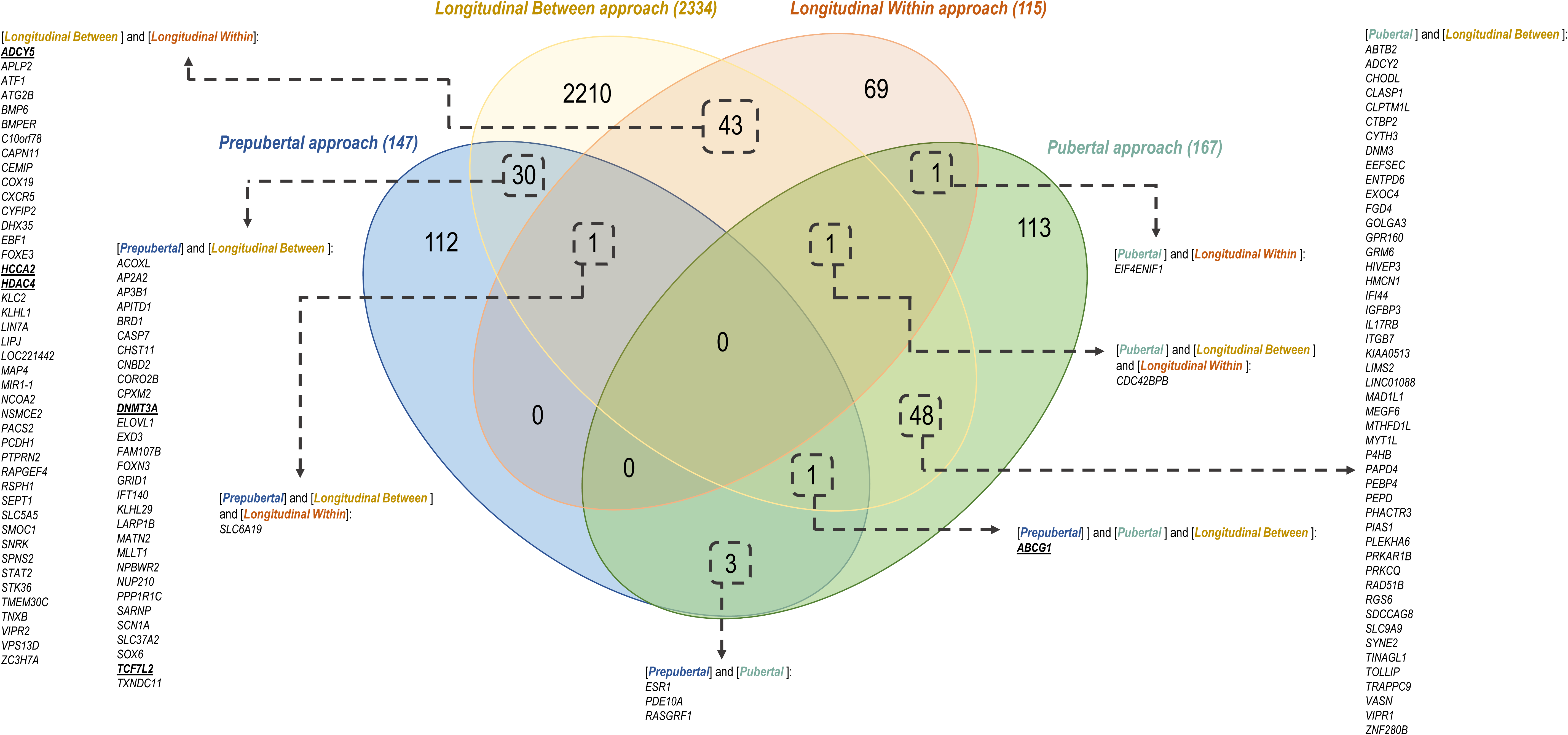
All significant associations showing a P-value < 0.005 with at least one trait in our continuous outcome DNA methylation analyses.

### 4. Association between gene expression and DNA methylation – A genome-wide eQTM analysis in human blood cells

With the intention of investigating the mechanistic relevance of identified IR epigenetics marks, RNA-seq analysis was performed in a subset of 44 children from the pubertal cross-sectional approach (**Figure 1**), which had also been included in the EWAS analysis. In this sub-population, we searched for cis-eQTMs in and around 10.000 bp of each transcript, and trans-eQTMs if the distance between the CpG and the transcript was higher than 10.000 bp. These analyses focused on the 267 CpGs from the ‘validation-list’ and 60,058 high-quality transcripts (whole-genome distributed) as input.

The cis-eQTM analysis identified 19 CpG-transcript pairs that met a P-value < 0.05, comprising 19 transcripts and 17 CpG sites (**Table 2A**). Methylation levels of CpG sites were both positively (45%) and negatively (55%) correlated with expression levels. Some genes reported in our regression analysis on obesity phenotypes were also revealed here as cis-eQTMs (highlighting *ABCG1*, *CEMIP*, *CNBD2*, *ESR1*, *FGD4*, *HMCN1* and *VASN*). Among them, only the *FGD4*, showed an FDR < 0.05. Identifed cis-eQTM CpGs for the genes *HMCN1*, *CASP7* and *VASN*, were annotated within enhancer regions according to the list of 450k enhancer predicted elements.

**Table 2.**
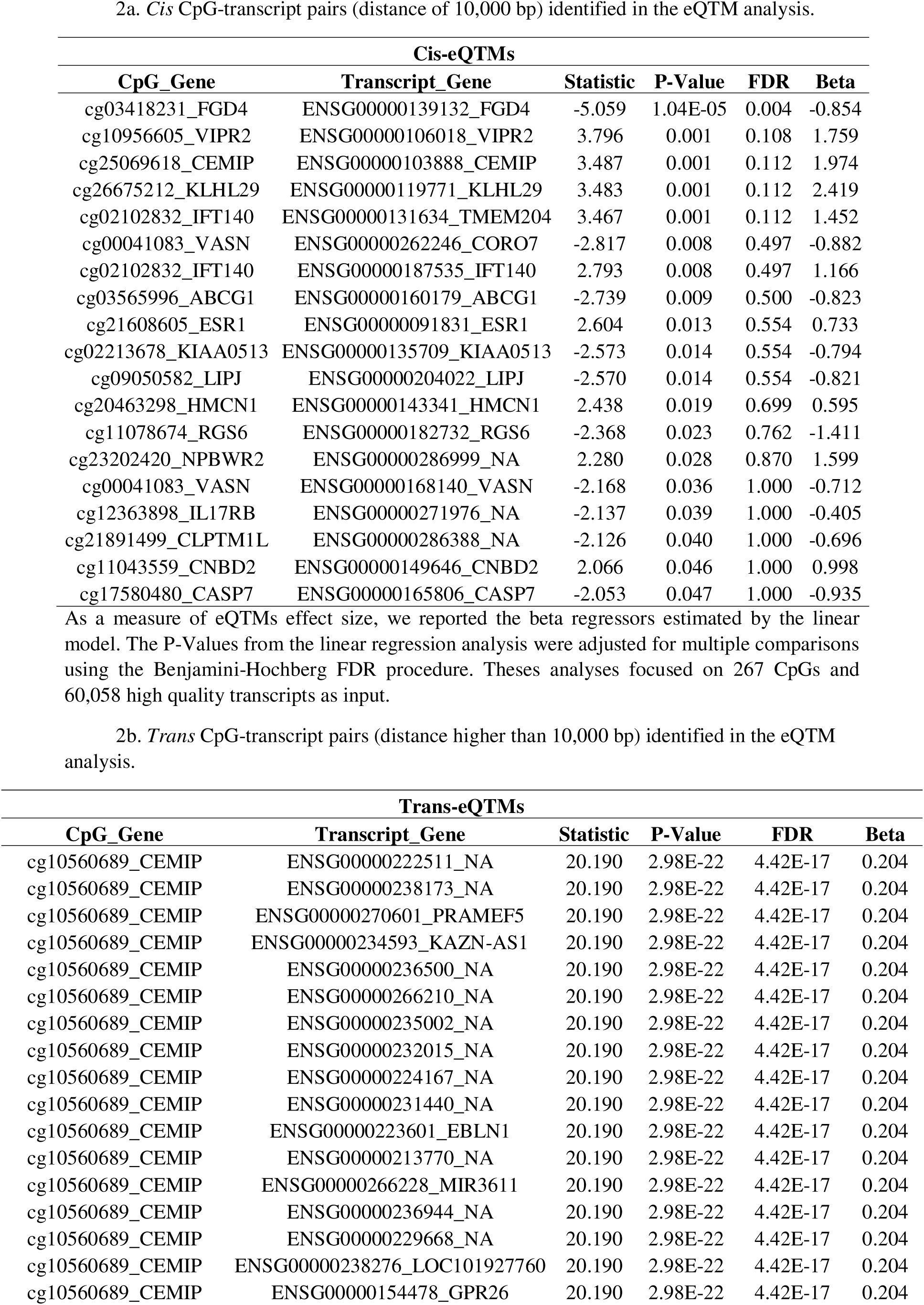
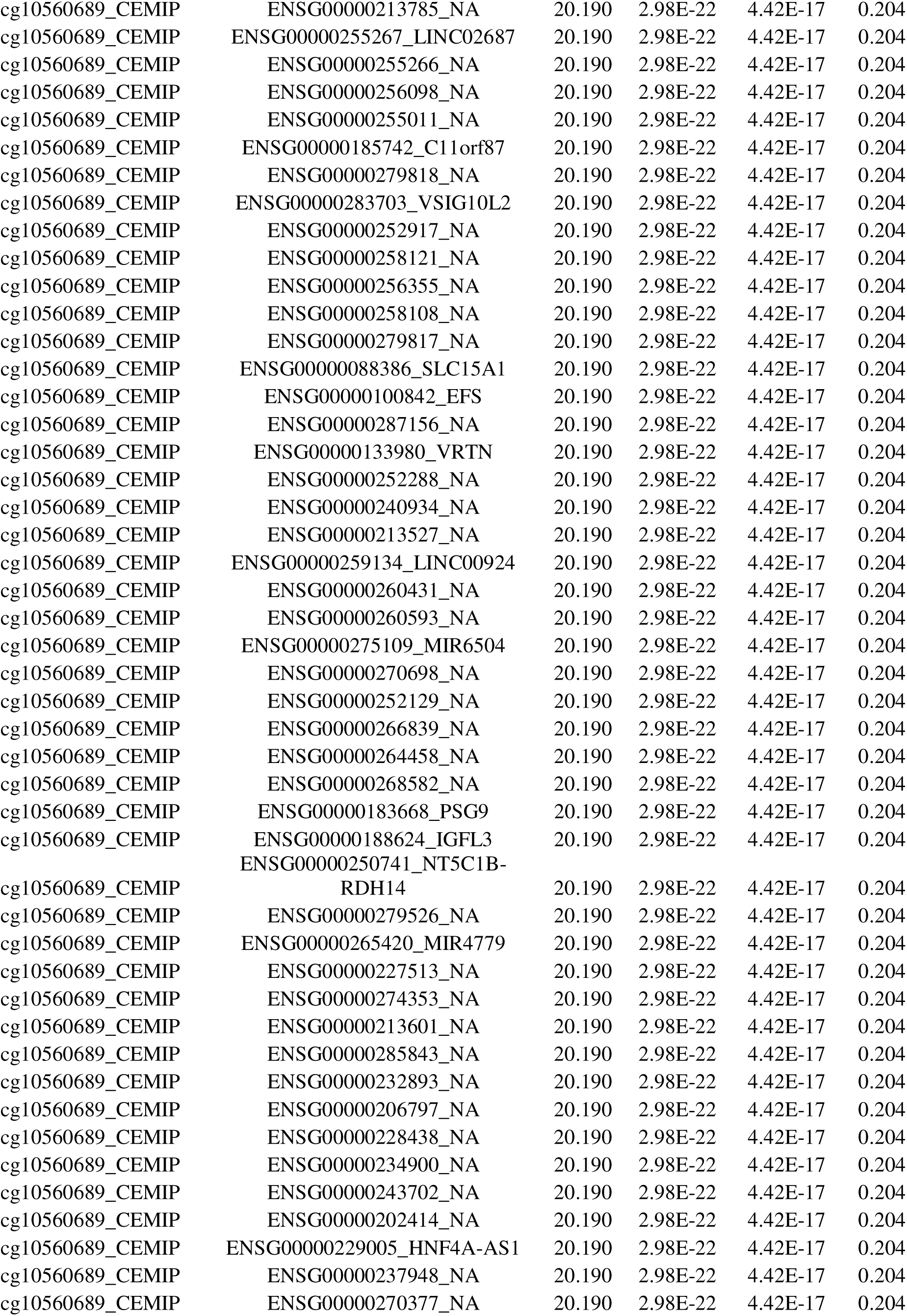
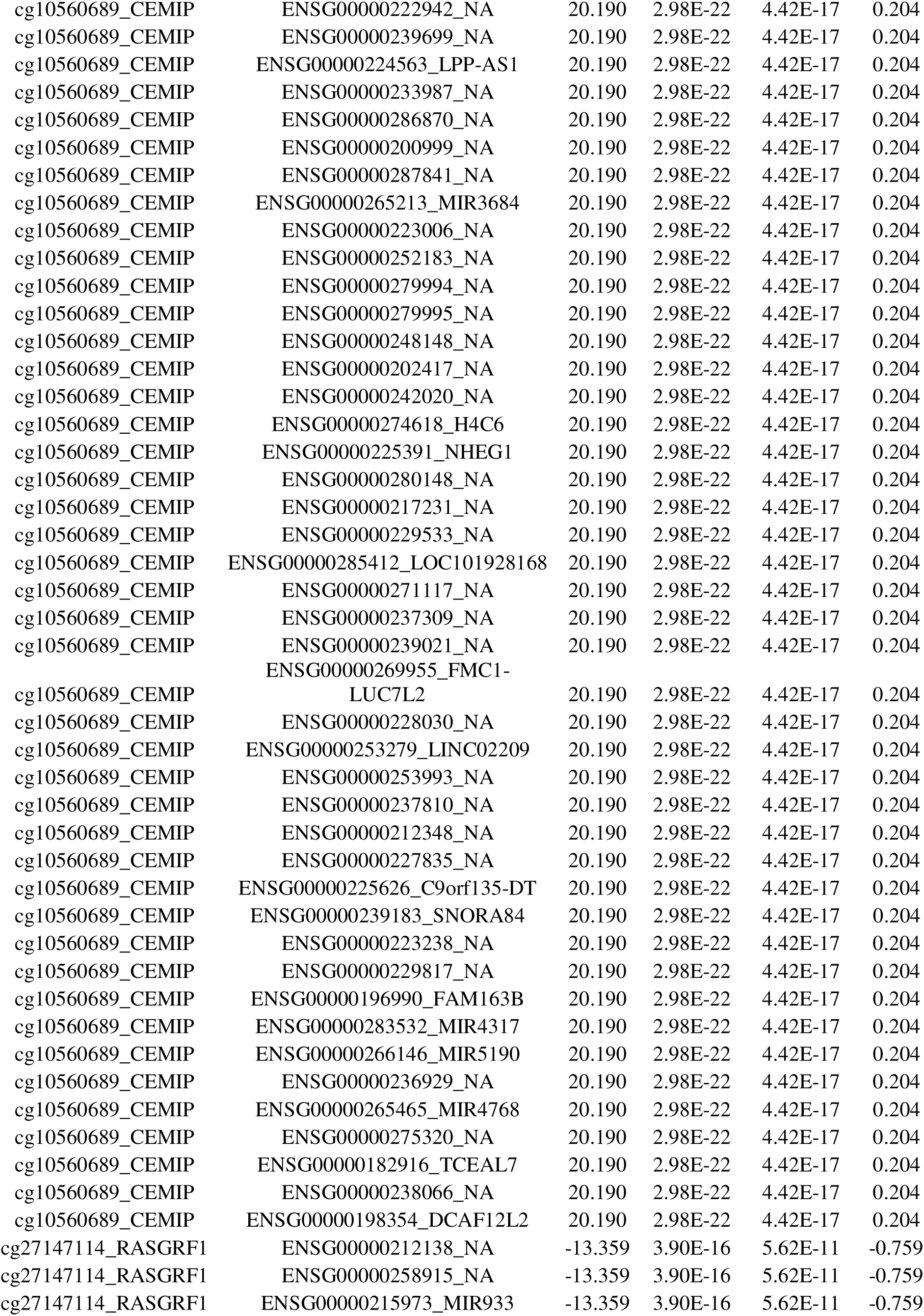
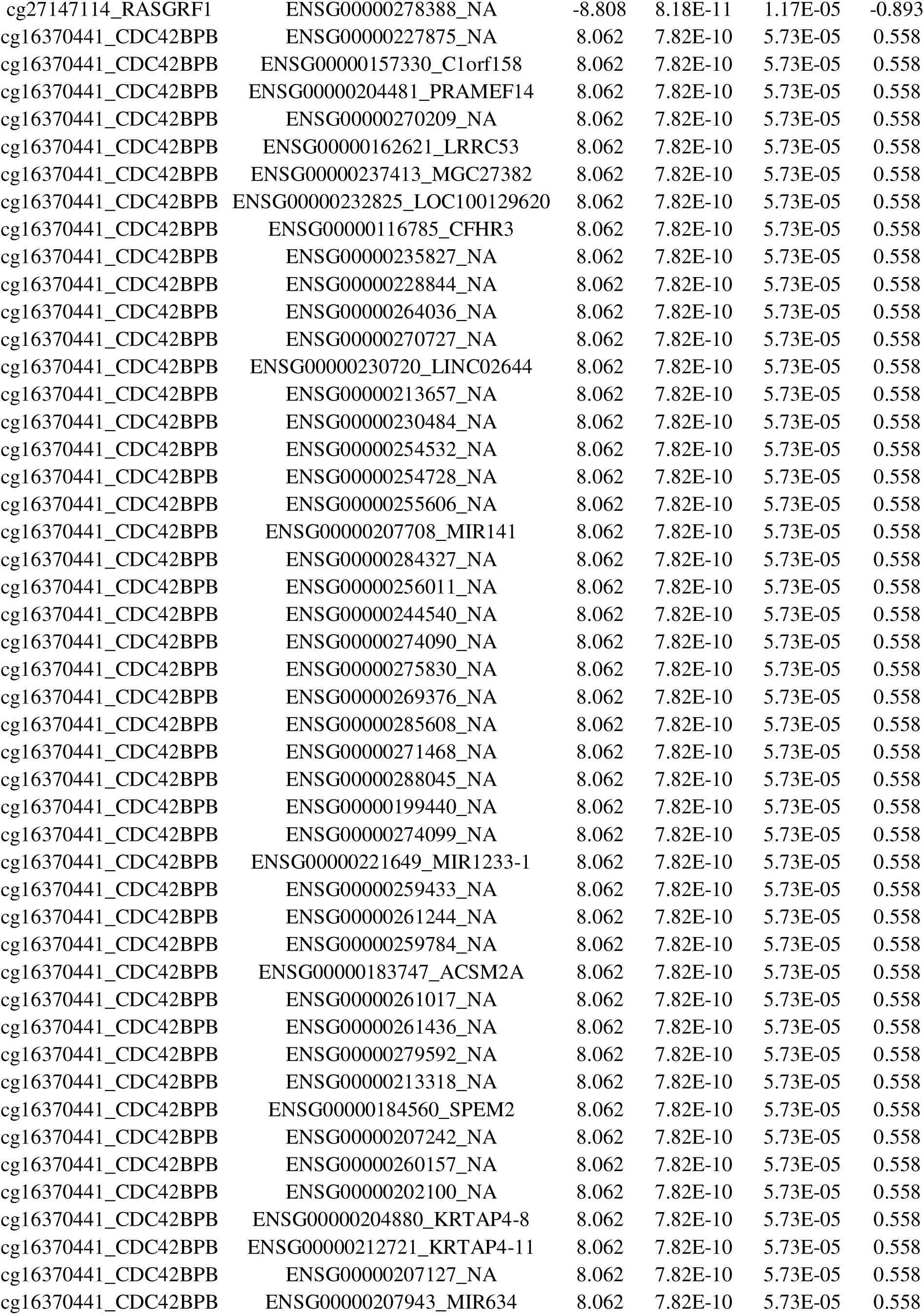
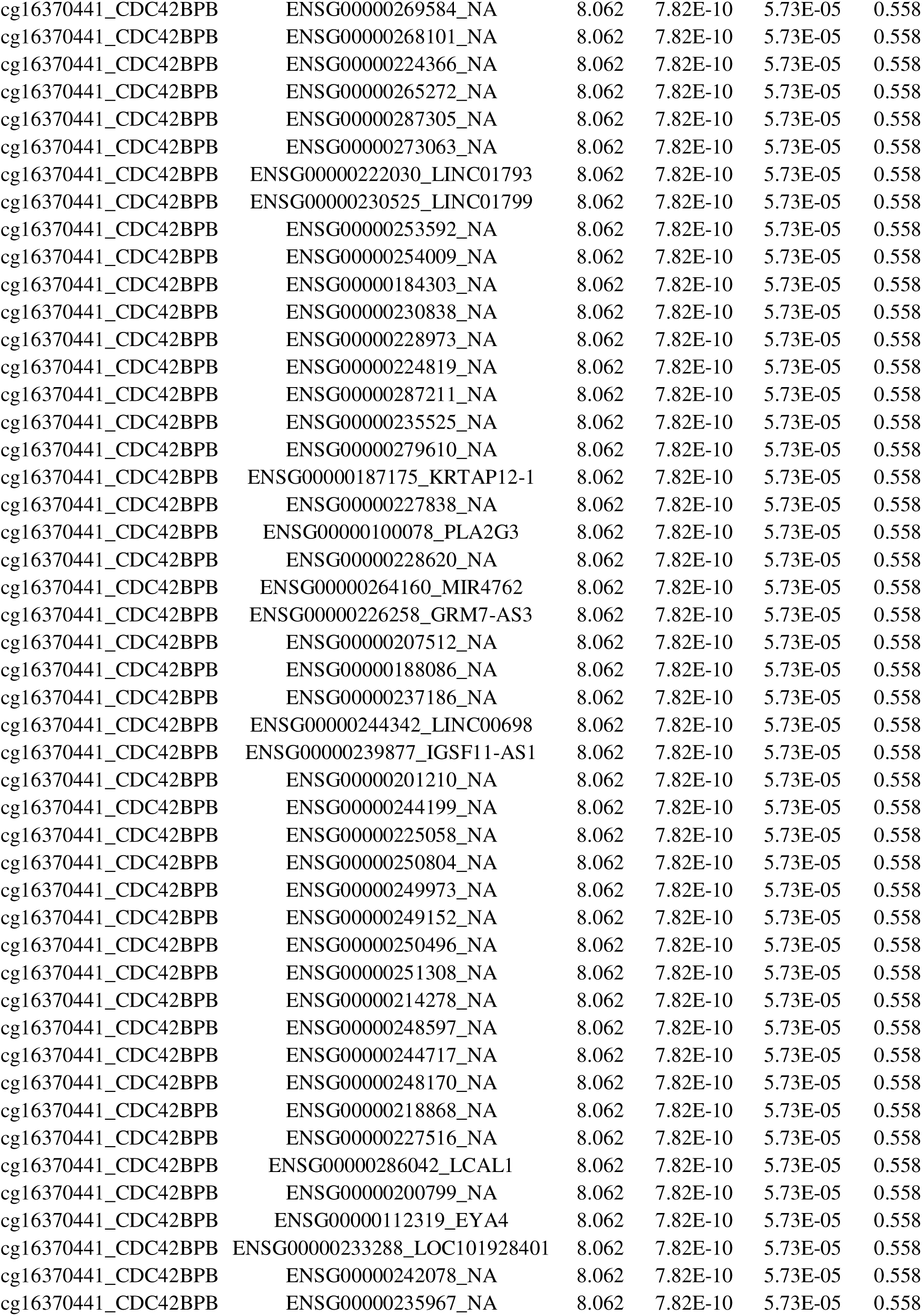
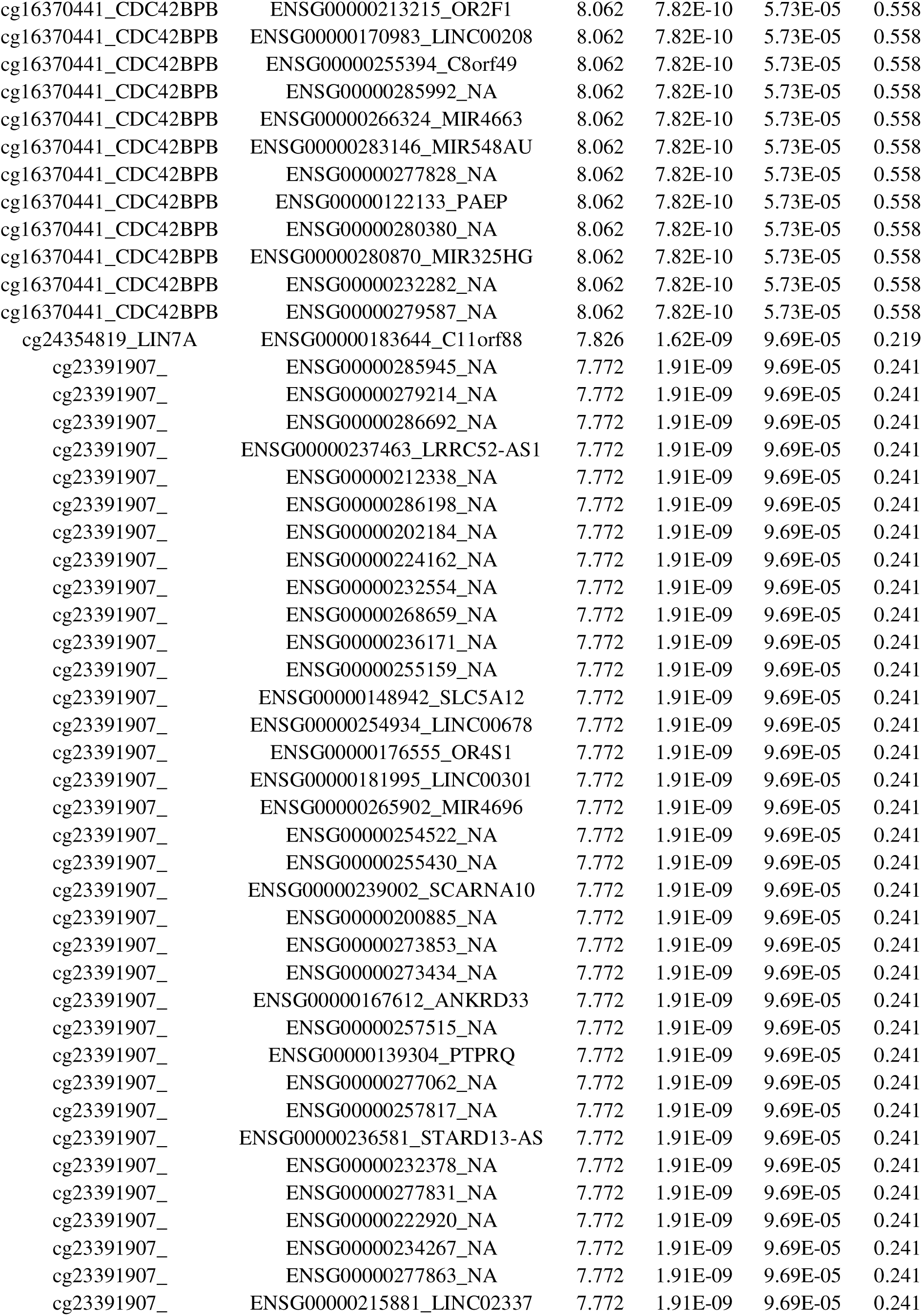
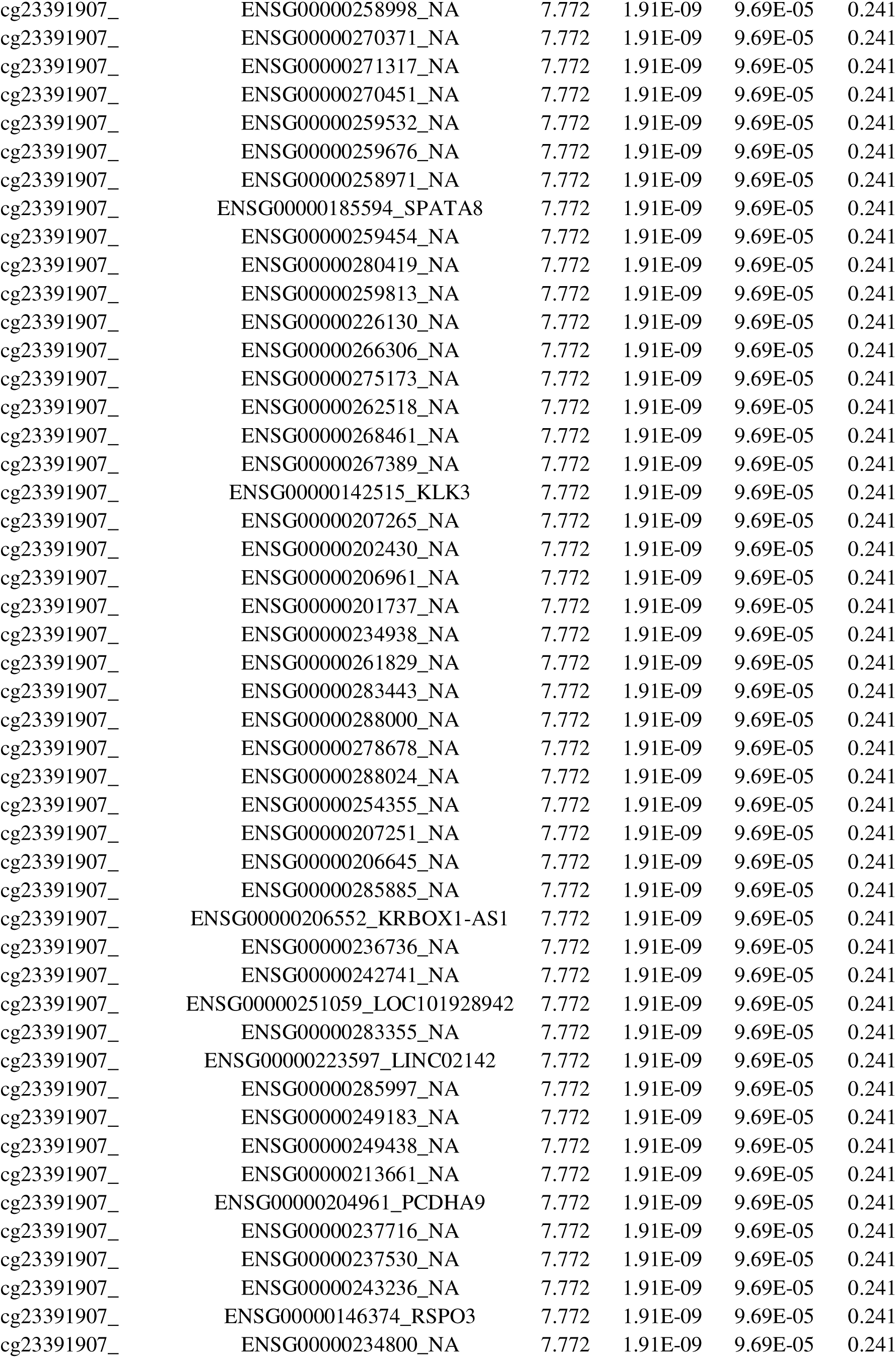
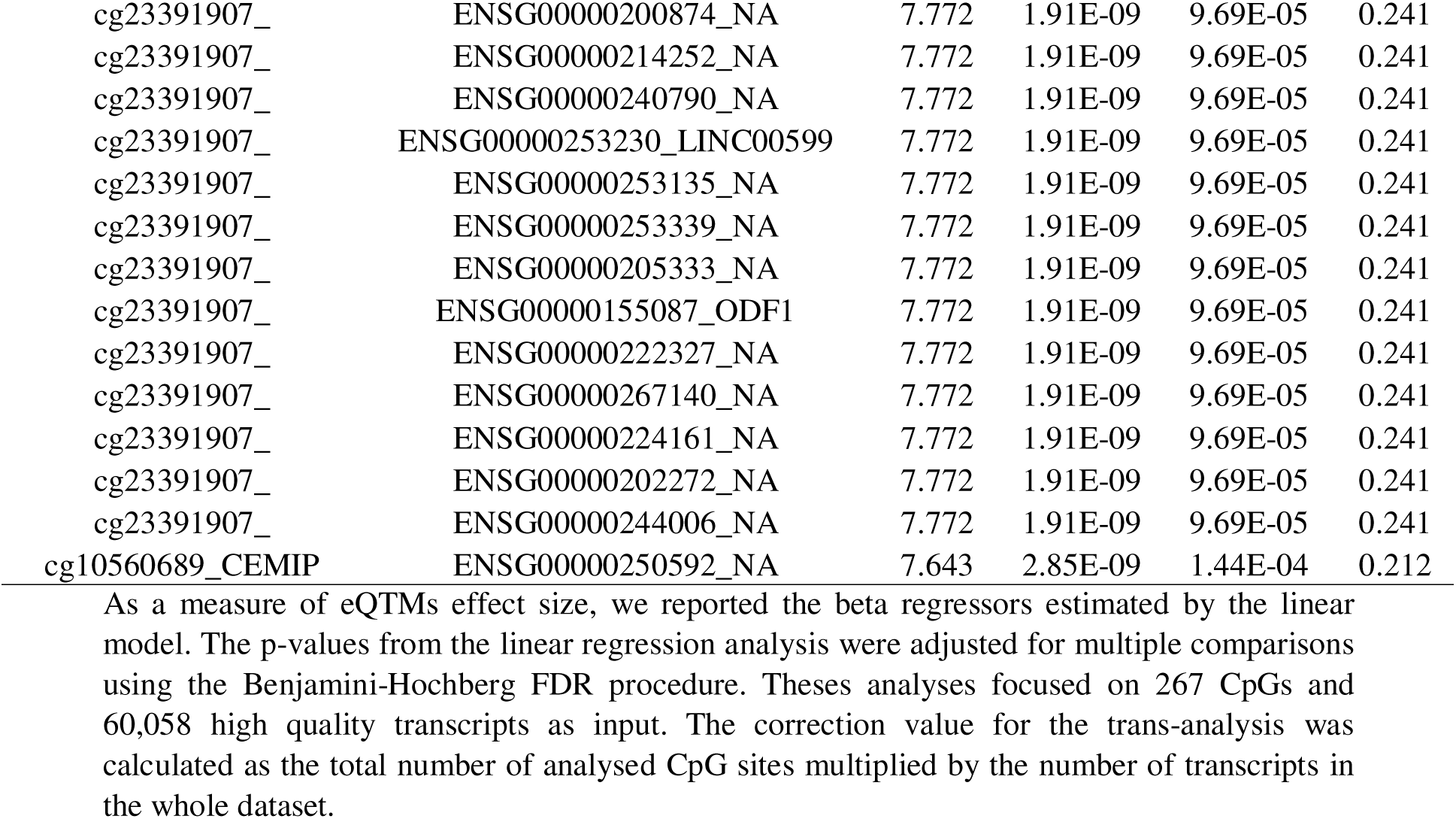
*Cis* CpG-transcript pairs (distance of 10,000 bp) identified in the eQTM analysis. Legend: As a measure of eQTMs effect size, we reported the beta regressors estimated by the linear model. The P-Values from the linear regression analysis were adjusted for multiple comparisons using the Benjamini-Hochberg FDR procedure. Theses analyses focused on 267 CpGs and 60,058 high quality transcripts as input.

The trans-eQTM analysis identified 317 CpG-transcript pairs that met an FDR < 1x10^-5^, comprising 317 transcripts, and 5 CpG sites mapping the genes *CDC42BPB*, *CEMIP*, *LIN7A* and *RASGRF1* (**Additional file 11**). Methylation levels of CpG sites were positively correlated with expression levels in the majority of trans-eQTM pairs (98.74%).

Identified cis-and trans-eQTMs were annotated according to the genomics regulatory elements they map (**Additional file 12**). The annotation consisted of two categories: 1) distance to a CpG island and 2) annotation to gene region. In terms of distance to CpG islands, we did not find differences in the annotation of eQTMs compared to the annotation derived from the whole list of CpGs in the EPIC array. On the contrary, in terms of annotation to gene regions, we found a higher proportion of CpGs mapping promoter regions (e.g., *TSS200* and untranslated regions [UTRs]) among the eQTMs in comparison to the whole list of CpGs in the EWAS EPIC array. Using the BIOS QTL browser [62, 63], we validated *in silico* some of the identified eQTMs. Particularly, we found evidences of blood cis-eQTMs for the same transcripts but distinct CpGs in the genes *ABCG1*, *CDC42BPB*, *ESR1*, *IFT140* and *LIN7A*.

Some identified cis-and trans-eQTM loci, like the *ABCG1*, *ESR1*, *FGD4*, *VIPR2*, *RGS6*, and *CEMIP*, mapped biological process GO-terms with relevance in obesity, puberty and metabolism; ‘activation of GTPase activity’, ‘antral ovarian follicle growth’, ‘cellular response to estrogen stimulus’, ‘G protein-coupled receptor signaling pathway’, ‘positive regulation of protein kinase C activity’, or ‘regulation of cholesterol esterification’.

### 5. Association between genetic variation and DNA methylation – A genome-wide mQTL analysis in human blood cells

At each cross-sectional approach (prepubertal and pubertal), mQTL analyses were conducted, revealing epigenetics regulating phenomena by which SNPs affect the methylation levels of a CpG. These analyses focused on the 267 CpGs from the ‘validation-list’ and 471,192 SNPs, whole-genome distributed, as input.

At the prepubertal stage, a total of 7 SNP-CpG pairs were found to be located in cis and 5 SNP-CpG pairs were located in trans (P-value < 0.05 and FDR < 0.005 respectively) (**Table 3**). Cis-mQTLs involved genes with special relevance to type 2 diabetes, such as the *ADCY5* or others previously highlighted in our pipeline, like *ESR1*. All but one SNP-CpG pair located in trans involved the gene *BRD1*. At the pubertal stage, a total of 10 SNP-CpG pairs were found to be located in cis and 10 SNP-CpG pairs were located in trans (P-value < 0.05 and FDR < 0.005 respectively) (**Table 4**). Cis-mQTLs for the loci *ADCY5*, *TINAGL1*, *GOLGA3* and *GRM6*, identified in the prepubertal approach, were also validated in the pubertal stage. The CpG mapping the *TINAGL1* was further annotated within an enhancer region. Two cis-mQTLs from the pubertal approach presented FDR < 0.05 (*MEGF6* and *SCN1A*). Another interesting gene, according to literature, and highlighted as cis-mQTL in the pubertal approach, is the *TNXB.* DNAm levels in this gene have been associated with an under nutrition status in adults [64] and previously reported as a mQTL in human pancreatic islets [39]. Using the BIOS QTL browser [62, 63], we validated *in silico* some of identified mQTLs. Particularly, we found evidence of blood cells cis-mQTLs for the genes (*ESR1*, *MEGF6* and *TNXB)*, although involving different CpGs and SNPs.

**Table 3a.** *Cis* SNP-CpG pairs (distance of 500 bp) identified in the mQTL analysis of the prepubertal stage. Legend: As a measure of mQTLs effect size, we reported the beta regressors estimated by the linear model. The p-values from the linear regression analysis were adjusted for multiple comparisons using the Benjamini-Hochberg FDR procedure.

| Cis-mQTLs |  |  |  |  |  |
| --- | --- | --- | --- | --- | --- |
| SNP | CpG_Gene | Statistic | P-Value | FDR | Beta |
| rs73186452 | cg00978808_ADCY5 | 3.391 | 9.99E-04 | 0.074 | 0.939 |
| GSA.rs114659838 | cg07504762_TINAGL1 | 3.074 | 0.003 | 0.089 | 1.672 |
| rs12282 | cg21561989_GOLGA3 | -2.980 | 0.004 | 0.089 | -0.592 |
| GSA.rs117733790 | cg23792592_MIR1-1 | 2.434 | 0.017 | 0.306 | 1.053 |
| GSA.rs35882398 | cg12001846_ESR1 | -2.351 | 0.021 | 0.306 | -1.302 |
| rs2067011 | cg10648542_GRM6 | 2.171 | 0.032 | 0.398 | 0.291 |
| GSA.rs114827188 | cg20329510_LIMS2 | -2.107 | 0.038 | 0.398 | -0.576 |

**Table 3b.**
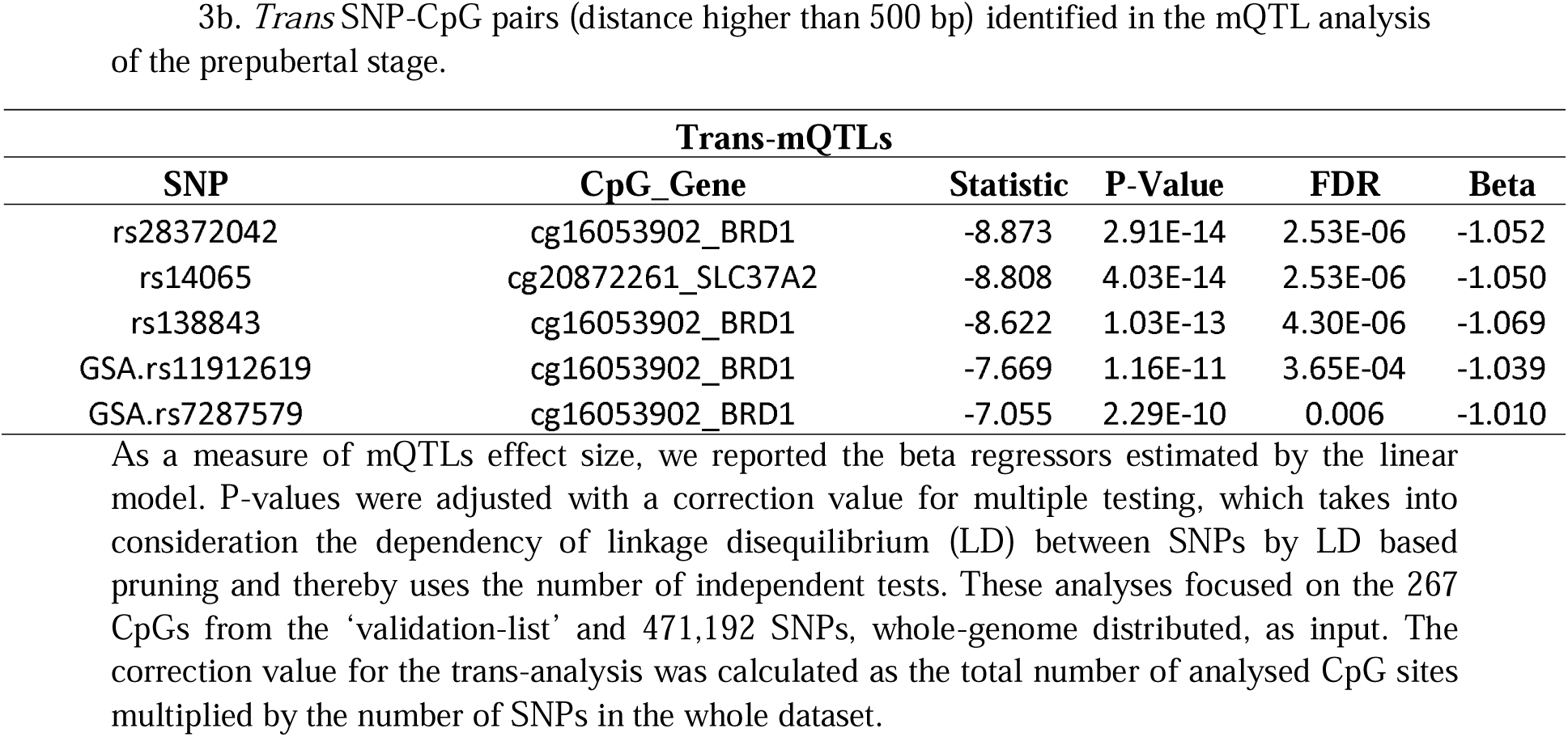
*Trans* SNP-CpG pairs (distance higher than 500 bp) identified in the mQTL analysis of the prepubertal stage. As a measure of mQTLs effect size, we reported the beta regressors estimated by the linear model. P-values were adjusted with a correction value for multiple testing, which takes into consideration the dependency of linkage disequilibrium (LD) between SNPs by LD based pruning and thereby uses the number of independent tests. These analyses focused on the 267 CpGs from the ‘validation-list’ and 471,192 SNPs, whole-genome distributed, as input. The correction value for the trans-analysis was calculated as the total number of analysed CpG sites multiplied by the number of SNPs in the whole dataset.

**Table 4a.** *Cis* SNP-CpG pairs (distance of 500 bp) identified in the mQTL analysis of the pubertal stage. Legend: As a measure of mQTLs effect size, we reported the beta regressors estimated by the linear model. The p-values from the linear regression analysis were adjusted for multiple comparisons using the Benjamini-Hochberg FDR procedure.

| Cis-mQTLs |  |  |  |  |  |
| --- | --- | --- | --- | --- | --- |
| SNP | CpG_Gene | statistic | P-Value | FDR | Beta |
| GSA.rs78267041 | cg00418943_MEGF6 | 3.750 | 0.00026329 | 0.019 | 1.583 |
| rs16851382 | cg15434576_SCN1A | -3.563 | 0.00051099 | 0.019 | -0.533 |
| rs12282 | cg21561989_GOLGA3 | -3.088 | 0.002 | 0.051 | -0.505 |
| rs7774197 | cg24252708_TNXB | -2.968 | 0.004 | 0.051 | -0.952 |
| GSA.rs114888185 | cg20320283_FOXE3 | -2.959 | 0.004 | 0.051 | -1.121 |
| GSA.rs114659838 | cg07504762_TINAGL1 | 2.918 | 0.004 | 0.051 | 1.258 |
| rs73186452 | cg00978808_ADCY5 | 2.447 | 0.016 | 0.166 | 0.679 |
| rs2067011 | cg10648542_GRM6 | 2.189 | 0.030 | 0.281 | 0.250 |
| rs3742476 | cg12913090_ATG2B | -2.131 | 0.035 | 0.287 | -0.254 |
| GSA.rs33932952 | cg25711726_SLC37A2 | 2.019 | 0.046 | 0.337 | 0.296 |

**Table 4b.** Tra*ns* SNP-CpG pairs (distance higher than 500 bp) identified in the mQTL analysis of the pubertal stage. Legend: As a measure of mQTLs effect size, we reported the beta regressors estimated by the linear model. P-values were adjusted with a correction value for multiple testing, which takes into consideration the dependency of linkage disequilibrium (LD) between SNPs by LD based pruning and thereby uses the number of independent tests. These analyses focused on the 267 CpGs from the ‘validation-list’ and 471,192 SNPs, whole-genome distributed, as input. The correction value for the trans-analysis was calculated as the total number of analysed CpG sites multiplied by the number of SNPs in the whole dataset.

| Trans-mQTLs |  |  |  |  |  |
| --- | --- | --- | --- | --- | --- |
| SNP | CpG_Gene | statistic | P-Value | FDR | Beta |
| GSA.rs11912619 | cg16053902_BRD1 | -11.263 | 4.87E-21 | 6.13E-13 | -1.189 |
| rs138843 | cg16053902_BRD1 | -11.116 | 1.14E-20 | 7.19E-13 | -1.148 |
| rs14065 | cg20872261_SLC37A2 | -10.820 | 6.34E-20 | 2.66E-12 | -0.953 |
| GSA.rs7287579 | cg16053902_BRD1 | -10.723 | 1.11E-19 | 3.50E-12 | -1.179 |
| rs7410612 | cg16053902_BRD1 | -10.361 | 8.97E-19 | 2.26E-11 | -1.121 |
| rs28372042 | cg16053902_BRD1 | -10.282 | 1.41E-18 | 2.97E-11 | -1.069 |
| rs1009321 | cg21561989_GOLGA3 | -7.104 | 6.79E-11 | 0.001 | -0.678 |
| GSA.rs761878 | cg16053902_BRD1 | -7.021 | 1.05E-10 | 0.002 | -1.077 |
| rs4477450 | cg20872261_SLC37A2 | -6.936 | 1.63E-10 | 0.002 | -0.732 |
| rs2824560 | cg20401955_CHODL | 6.807 | 3.17E-10 | 0.004 | 0.678 |

### 6. Serum protein levels of vasorin are associated with IR and obesity in the pubertal stage

To further investigate the role of one of the most promising biomarkers identified for IR, we measured VASN serum protein levels in the cohort. Descriptive statistics for experimental groups reported lower levels of VASN protein significantly associated with IR and obesity in the pubertal stage of the children (P=0.007) (Additional file 3). Moreover, in the longitudinal approach (N = 90), groups maintaining or developing IR with the onset of puberty, G4 and G5, presented the lowest increases in VASN levels (P = 0.06) (Additional file 13). At the opposite, insulin-sensitive groups such as G1 and G3 showed a pronounced increase in VASN levels.

With the aim of functionally validate our EWAS and eQTM findings for *VASN* in the pubertal stage, we also studied the correlation between *VASN* DNAm and VASN serum protein levels, as well as between *VASN* mRNA and VASN serum protein levels.

Interestingly, in the 130 pubertal children, a suggestive trend was reported for the correlation between DNAm at the cg00041083 and VASN protein levels after adjusting for confounders such as age, sex, origin and BMI Z-Score (P = 0.09) (Additional file 14); higher DNAm levels related to lower protein levels. Contrarily, in the 44 pubertal children with available RNAseq data, we did not find a significant correlation between mRNA levels at the ENSG00000168140 and VASN serum levels (P = 0.34).

## Discussion

The current large-scale integrative molecular analysis identifies novel blood multi-omics signatures such as DNAm marks, eQTMs and mQTLs, underlying the development, amelioration and worsening of IR in children with obesity during puberty. Functional enrichment analysis revealed that identified loci participate in systemic metabolic pathways and sexual maturation processes with relevance to the pathogenesis of IR. Additional analyses on cardiometabolic and inflammatory phenotypes show that blood DNAm patterns of some identified loci are further associated, beyond IR, with an overall risky-cardiometabolic profile in children. To our knowledge, this is the first longitudinal multi-omics approach characterizing molecular blood alterations for IR and obesity during the metabolically critical period of puberty. With our results, we propose novel and promising biomarkers with predictive utility for the identification of children with obesity at high risk of developing IR and metabolic alterations. Likewise, we also aid insights into the molecular and functional mechanisms linking epigenetics alterations and the IR phenotype in obesity.

Our EWAS analysis on IR identified 4,281 associated unique CpG sites, from which 2,981 further presented an FDR < 0.05 (linked to 2,632 and 1,899 genes, respectively) (**Figure 1**). Among them, we selected only those loci presenting significant associations in at least two of our statistical approaches (**Figure 3**). The resulting list was composed of 267 IR-associated unique CpGs mapping 128 genes, from which 130 CpGs (mapping 91 genes) presented an FDR < 0.05 (**Figure 4 and Additional file 7**). Among our top significant results (**Table 1** and **Additional file 7**), there were new and promising regions never reported as epigenetics marks of IR (e.g., *CDC42BPB, ESR1, HMCN1*, *PRKAR1B*, *SNRK* and *VASN*, among others). From them, DNAm levels of the *ESR1* showed association with IR not only in the pubertal but also in the prepubertal stage (**Figure 3**), indicating that they might accompany IR already from early childhood. The rest of them otherwise showed association with IR in our longitudinal and pubertal approaches, resembling marks associated with IR in the context of puberty. On these and the rest loci from our ‘validation-list’, associations with a bulk of cardiometabolic phenotypes other than IR were also investigated (**Figure 5**). As a novelty, these confounding-adjusted analyses allowed us to distinguish between regions in which the initial IR-association is direct (e.g., *ABCG1* and *VASN*), or rather derives from an indirect or secondary association with anthropometry and obesity traits (e.g., *CNBD2* and *FGD4*), or with inflammation (e.g., *CDC42BPB*). Interestingly, the functional enrichment analysis of reported CpGs indicated that identified loci participate in systemic metabolic pathways, inflammatory and sexual maturation processes with relevance to the pathogenesis of IR. Among them, the terms related to the synthesis and secretion of sexual hormones outline the importance of puberty and its hormonal and biochemical changes as plausible contributors to the development and worsening of obesity IR.

Beyond the new targets identified, we also found genes whose methylation levels have been previously and repeatedly associated with adult type 2 diabetes and obesity in the literature (e.g., *ABCG1, ADCY5, CPT1A, FTO, HCCA2, HDAC4, HIF3A, IGF-1, KCNQ1, PPARG,* and *TCF7L2,* among others) [13, 23]. Therefore, our results remark the role of IR as an important pathophysiological mechanism linking obesity and cardiometabolic comorbidities, and reinforce the fact that epigenetics marks of IR may have utility as predictive markers of future disease outcomes. In addition to type 2 diabetes, our associations also highlighted loci specifically associated with adult IR in the literature (e.g., *COL18A1, CTNND2, CXCL1, DNMT3A, GRB10, HDAC4, LAT, PAX6, SH3RF3* and *SIRT2*) [18–22]. This is important since literature IR studies had mostly focused on studying the relationship between the methylation levels of candidate genes and the HOMA-IR [22], and EWAS on IR are still scarce [14–16, 22, 65] (with barely one study conducted in children) [66]. The fact of validating here previously known adult epigenetics marks of IR may indicate that the DNA methylation patterns of IR are established early, under the influence of childhood or puberty obesogenic environments, and remain stable throughout adulthood.

In order to elucidate the molecular mechanisms behind identified IR epigenetics marks, we integrated our EWAS data along with other omics sources in the same cohort (GWAS and RNAseq data). As a result, we reported that some of the identified loci might be participating in phenomena that alter gene expression levels (eQTMs) while others could be explained by the existence of SNPs (mQTLs) (**Figure 1**).

Regarding eQTMs phenomena, our analysis reported that some of the most promising regions identified could exert their effects of IR through a modification, either up-or down-regulating, the expression of target transcripts *in situ* (*cis*-eQTMs), or at long distances from their occurrence (*trans*-eQTMs) (**Table 2**). Among identified *cis*-eQTM phenomena, we can highlight previously well-known eQTM loci (like the *ABCG1*) [23] but also some of our promising new markers (*CDC42BPB, ESR1, HMCN1* and *VASN*). Interestingly, most identified CpGs mapped into genomics regulatory elements (e.g., enhancers, transcription start sites or UTR), reinforcing their role as plausible gene expression controllers. To date, this is the first study integrating RNAseq and EWAS data in the blood of pubertal insulin-resistant children with obesity. Previously, a recent study investigated the existence of eQTMs phenomena in the adipose tissue of African American adult women with IR [17]. Although with no overlapping regions between their and our approach, our findings reinforce the idea that DNAm-mediated regulation of gene expression could be implicated into the pathogenesis of IR.

Previous studies have shown that DNAm alterations at the cg06500161 of *ABCG1* strongly correlate with glucose metabolism dysfunction and diabetes in adults [14, 65, 67]. Many of these studies have also evidenced that these DNAm alterations elicit effects on *ABCG1* gene expression through eQTM interactions. Moreover, the connection between *ABCG1* and diabetes-related traits and dyslipidemia has been supported by animal and human studies [14, 65, 67]. The protein encoded by *ABCG1* is a member of the superfamily of ATP-binding cassette (ABC) transporters. ABC proteins transport various molecules across extra-and intracellular membranes. More specifically, ABCG1 is involved in macrophage cholesterol and phospholipid transport and may regulate cellular lipid homeostasis in other cell types. Although a bulk of insights had been reported in adults, our results are the first evidence of such relationships in children with obesity and IR. Thus, we contribute to the body of evidence supporting the role of lipid metabolism, specifically of ABC transporters, in IR and highlight the interest of *ABCG1* as a potential non-invasive biomarker for future glucose metabolism complications.

Here, mQTL analyses were also conducted revealing epigenetics regulating phenomena, by which SNPs affect the methylation levels of CpGs (**Figure 1**). For this approach, we counted on the EWAS data from both the prepubertal and pubertal stages, which allowed us to look for replicated mQTL phenomena across time points. Among results, we again reported some previously known diabetes loci (such is the case of *ADCY5*) [68, 69] but also interesting new IR epigenetics marks (*ESR1*), for which previous mQTL evidence had been reported in the literature [13]. These phenomena could be, therefore, the molecular explanation for some epigenetics IR early-life marks for which the environment is not the causal mediator.

*ESR1* is a non-imprinted gene that encodes the estrogen receptor-α (ER-α), a transcription factor involved in the regulation of energy homeostasis [70]. In females, estrogens maintain energy homeostasis via ERα by suppressing energy intake and lipogenesis, enhancing energy expenditure [71] and ameliorating insulin secretion and sensitivity [72]. In males, however, testosterone is converted to estrogen and maintains fuel homeostasis via ERα and androgen receptors, which share related functions to suppress adipose tissue accumulation and improve insulin sensitivity. Conversely, the lowering in estrogens levels observed in postmenopausal women provoke IR and increase the risk of type 2 diabetes [70]. Although no previous evidence of an association between DNAm levels and IR has been reported for the *ESR1* in the literature, dynamic changes in the DNAm of this region have been previously associated with aging [73]. It is noteworthy to see how the *ESR1*, which had not been previously evidenced as an epigenetic marker of IR, appears as a significant locus in all the approaches of our study (EWAS on IR, association with cardiometabolic traits, *cis-* eQTMs and *cis*-mQTLs). From this, we can conclude several things; 1) DNAm alterations in the *ESR1* locus during puberty could be an important contributor to the appearance and worsening of IR in children with obesity, 2) these alterations could exert their effects on the phenotype through the alteration of *ESR1* gene expression levels, and 3) there could also be some from-birth predisposing SNPs favouring the alteration of DNAm levels in the region. The evidenced mQTL phenomenon of the region agrees with the fact of *ESR1* appearing as a significant epigenetic marker of IR in both prepubertal and pubertal stages in our study. Considering our results and the implication of the estrogen axis into the context of IR and puberty, we propose that *ESR1* could be a promising epigenetic target to prevent age-related metabolic disorders associated with obesity.

Besides the *ESR1*, the most promising and novel biomarker identified from our approach is the *VASN*, which was reported as a top association from the ‘validation-list’ in the EWAS on IR (**Figure 4** and **table 1**), as well as a participant of a *cis*-eQTM phenomenon. Particularly, we report for the first time that both higher blood *VASN* DNAm levels and lower serum protein concentrations are strongly associated with IR in the pubertal stage in children with obesity. Although our eQTM analysis showed that the higher DNAm of *VASN* is associated with lower mRNA *VASN* levels in our children, we could not validate the results with an association between mRNA *VASN* levels and VASN serum levels. This is not surprising otherwise since the elevated VASN serum levels associated with IR are a systemic finding that could derive from many other tissues than blood cells. Moreover, the population sample size with RNAseq data and VASN serum levels measured was small (barely 40 subjects). VASN is a type I transmembrane protein (SLIT-like 2), highly expressed in smooth muscle cells and with reported expression in adipocytes [74, 75]. VASN was originally found to play a role in vascular injury repair and angiogenesis, and is a potential biomarker for hepatocarcinoma [75]. Mechanistically, VASN directly binds to the transforming growth factor (TGF-B) and attenuates TGF-B signaling *in vitro*. A recent study has also shown that hypoxia increases Notch signaling in glioma-like cells through the induction of VASN and the *hypoxia-inducible factor-1* (HIF1)/STAT3), thus describing a possible action mechanism of VASN. However, the relationship between VASN, obesity and IR remains unknown. Our main hypothesis is that *VASN* could play an important role in obesity as a potential biomarker of IR and/or a predictor of future development of type 2 diabetes in children.

The main limitations of our study are, on the one hand, the low number of participants included in our populations. On the other hand, it is the fact that our findings are based on data from blood, which was the only accessible tissue, and may not be representative of other metabolically relevant organs such as live and adipose and muscle tissues. In this regard, there is a trend pointing to a correlation between the global state of methylation in blood and adipose tissue [76]. This correlation might be explained by the abundant presence of white cells in both tissues and suggests that buffy coat might be a valid indicator of what happens at the methylation level in adipose tissue, especially for the case of inflammatory and immune system-related aspects. Another possible source of bias would be the difference in time elapsed between the two measurements (prepubertal and pubertal times) between the different participants.

The main strengths of our study are the high significance of our associations (many of them passing multiple-test correction thresholds) as well as the multi-omics design, from which we validate our top associations in a multi-omics dimensional space. Likewise, another positive point is the future pubertal study design, which strengthens the statistical robustness of our reports. Finally, it is a fact of being the first longitudinal multi-omics approach characterizing molecular blood alterations for IR and obesity during the metabolically critical period of puberty.

## Conclusions

With our results, we propose novel and promising biomarkers of IR and metabolic alterations in children with obesity (*ABCG1, CDC42BPB, ESR1, HMCN1*, *PRKAR1B*, *SNRK* and *VASN*, among others). Thanks to our multi-omics design, we also aid insights into the molecular and functional mechanisms linking epigenetics alterations and the IR phenotype in obesity (mQTLs and eQTMs). If validated in other cohorts and longitudinal designs, our identified loci could serve as predictive non-invasive biomarkers for reducing the high rates of mortality and morbidity associated with obesity. Especially for genes with a promising but unknown role in the development of IR in the adipose tissue, such is the case of *VASN*, additional *in vitro* and *in vivo* functional analyses should be conducted in the near future.

## Data Availability

All data produced in the present study are available upon reasonable request to the authors

## Abbreviations

CVD: Cardiovascular disease
DMS: Differentially methylated site
DNAm: DNA methylation
EWAS: Epigenome-Wide Association Studies
eQTMs: Expression quantitative trait methylations
FDR: False discovery rate
GWAS: Genome-Wide Association Studies
HWE: Hardy-Weinberg equilibrium
IR: Insulin resistance
mQTLs: Methylation quantitative trait loci
MAF: Minor allele frequency
MLR: Multiple linear regression
SNPs: Single nucleotide polymorphisms

## Acknowledgments

The authors would like to thank the children and parents who participated in the study. This work was supported by the Plan Nacional de Investigación Científica, Desarrollo e Innovación Tecnológica (I + D + I), Instituto de Salud Carlos III-Health Research Funding (FONDOS FEDER) (PI16/00871, PI20/00563 and PI20/00711); the Regional Government of Andalusia (“Plan Andaluz de investigación, desarrollo e innovación (2018), P18-RT-2248); Redes temáticas de investigación cooperativa RETIC (Red SAMID RD12/0026/0015) and the Mapfre Foundation. The authors also acknowledge Instituto de Salud Carlos III for personal funding: Contratos i-PFIS: doctorados IIS-empresa en ciencias y tecnologías de la salud de la convocatoria 2017 de la Acción Estratégica en Salud 2013–2016 (IFI17/00048).

## ’Availability of data and materials’

The datasets supporting the conclusions of this article are available in the EGA and GEO repositories.

## Additional material

**Additional file 1.** File format: .xls. Descriptive statistics for the longitudinal population (N=90).

**Additional file 2.** File format: .xls. Descriptive statistics for the cross-sectional prepubertal population (N=99).

**Additional file 3.** File format: .xls. Descriptive statistics for the cross-sectional prepubertal population (N=130).

**Additional file 4.** File format: .png. PCA plot grouping the technical replicates (samples) among runs of RNAseq analysis.

**Additional file 5.** File format: .xls. Whole list of significant associations derived from the EWAS on insulin resistance.

**Additional file 6.** File format: .xls. KEGG-pathway enrichment analysis for the whole list of significant associations derived from the EWAS on insulin resistance.

**Additional file 7.** File format: .xls. Whole list of overlapping findings (at loci level) between the statistical approaches of the EWAS on insulin resistance. These findings constitute the so called ‘validation-list’.

**Additional file 8.** File format: .xls. KEGG-pathway enrichment analysis for the CpGs in the ‘validation-list’.

**Additional file 9.** File format: .xls. GO-terms enrichment analysis for the CpGs in the ‘validation-list’.

**Additional file 10.** File format: .pdf. All significant associations showing a P-value < 0.05 with at least one trait in our continuous outcome DNA methylation analyses.

**Additional file 11.** File format: .word. *Trans* CpG-transcript pairs (distance higher than 10,000 bp) identified in the eQTM analysis. Legend: As a measure of eQTMs effect size, we reported the beta regressors estimated by the linear model. The p-values from the linear regression analysis were adjusted for multiple comparisons using the Benjamini-Hochberg FDR procedure. These analyses focused on 267 CpGs and 60,058 high-quality transcripts as input. The correction value for the trans-analysis was calculated as the total number of analyzed CpG sites multiplied by the number of transcripts in the whole dataset.

**Additional file 12.** File format: .pdf. Proportions of identified cis-and trans-eQTMs according to the genomics regulatory elements they map.

**Additional file 13.** File format: .pdf. Longitudinal trajectories of VASN Serum levels in the experimental groups of our longitudinal approach (before and after puberty).

**Additional file 14.** File format: .pdf. Regression plot between *VASN* DNAm (cg00041083) and VASN Serum levels.

